# Neural adaptation of cingulate and insular activity during delayed fear extinction: A replicable pattern across assessment sites and repeated measurements

**DOI:** 10.1101/2020.10.26.20218206

**Authors:** Isabelle C. Ridderbusch, Adrian Wroblewski, Yunbo Yang, Jan Richter, Maike Hollandt, Alfons O. Hamm, Hans-Ulrich Wittchen, Andreas Ströhle, Volker Arolt, Jürgen Margraf, Ulrike Lueken, Martin J. Herrmann, Tilo Kircher, Benjamin Straube

**Affiliations:** Department of Psychiatry and Psychotherapy & Center for Mind, Brain and Behavior -CMBB, Philipps-University Marburg; Institute for Psychology, University of Greifswald; Institute for Clinical Psychology and Psychotherapie, Technichal University of Dresden; Department of Psychiatry and Psychotherapy, Ludwig-Maximilians-University (LMU) Muenchen; Klinik für Psychiatrie und Psychotherapie, Campus Charité Mitte, Charité - Universitätsmedizin Berlin, corporate member of Freie Universität Berlin, Humboldt-Universität zu Berlin and Berliner Institut für Gesundheitsforschung; Department of Psychiatry and Psychotherapy, University Hospital Muenster; Chair in Psychiatry and Psychotherapy, Ruhr University Bochum (RUB); Center for Mental Health, Department of Psychiatry, Psychosomatics, and Psychotherapy, University Hospital Wuerzburg; Department of Psychology, Humboldt-Universität zu Berlin

**Keywords:** fear conditioning, extinction, fMRI

## Abstract

Adapting threat-related memories towards changing environments is a fundamental ability of organisms. One central process of fear reduction is suggested to be extinction learning, experimentally modeled by extinction training that is repeated exposure to a previously conditioned stimulus (CS) without providing the expected negative consequence (unconditioned stimulus, US). Although extinction training is well investigated, evidence regarding process-related changes in neural activation over time is still missing. Using optimized delayed extinction training in a multicentric trial we tested whether: 1) extinction training elicited decreasing CS-specific neural activation and subjective ratings, 2) extinguished conditioned fear would return after presentation of the US (reinstatement), and 3) results are comparable across different assessment sites and repeated measures. We included 100 healthy subjects (measured twice, 13-week-interval) from six sites. 24h after fear acquisition training, extinction training, including a reinstatement test, was applied during fMRI. Alongside, participants had to rate subjective US-expectancy, arousal and valence. In the course of the extinction training, we found decreasing neural activation in the insula and cingulate cortex as well as decreasing US-expectancy, arousal and negative valence towards CS+. Re-exposure to the US after extinction training was associated with a temporary increase in neural activation in the anterior cingulate cortex (exploratory analysis) and changes in US-expectancy and arousal ratings. While ICCs-values were low, findings from small groups suggest highly consistent effects across time-points and sites. Therefore, this delayed extinction fMRI-paradigm provides a solid basis for the investigation of differences in neural fear-related mechanisms as a function of anxiety-pathology and exposure-based treatment.

**Clinical Trials Registration:** Registry names: Deutsches Register Klinischer Studien (DRKS) – German Clinical Trails Register

ClinicalTrials.gov

Registration ID-numbers: DRKS00008743

DRKS00009687

ClinicalTrials.gov Identifier: NCT02605668

URLs: https://www.drks.de/drks_web/navigate.do?navigationId=trial.HTML&TRIAL_ID=DRKS00008743

https://www.drks.de/drks_web/navigate.do?navigationId=trial.HTML&TRIAL_ID=DRKS00009687

https://clinicaltrials.gov/ct2/show/NCT02605668

## 1. Introduction

Extinction training gets an individual to learn inhibiting a previously conditioned response (CR) to a fear conditioned stimulus (Lonsdorf et al., 2017). It is neither forgetting nor “unlearning” (Vervliet et al., 2013) but a new learning process competing with a previously established stimulus-reaction relation. Extinction learning is adaptive adjusting one’s reactions to changing circumstances and learning that a prior threat signal no longer indicates danger is particularly evolutionary vital.

Extinction learning as a re-evaluation of threat prediction is limited in patients with anxiety disorders (Duits et al., 2015), that affect up to 25% of the general population worldwide (Remes et al., 2016) and cause substantial individual and societal burden (Gustavsson et al., 2011; Wittchen et al., 2011). Currently the most effective treatment of anxiety disorders is exposure-based cognitive-behavioral therapy (CBT) (Bandelow et al., 2014; National Institute for Health and Clinical Excellence (NICE), 2011, 2013). Extinction learning is suggested to be one central process of fear reduction in exposure-based CBT (Milad & Quirk, 2012; Vervliet et al., 2013; Heinig et al., 2017; Richter et al., 2017). During exposure sessions, patients are confronted repeatedly with situations that evoke their pathological anxieties to re-learn that these “threat” signals do not indicate actual danger, which over time and repetition reduces pathological fear responses.

In the experimental laboratory model, the effects of fear extinction training are investigated subsequently to fear acquisition training in which a neutral stimulus (conditioned stimulus, CS) is repeatedly paired with an aversive stimulus (unconditioned stimulus, US) (Pavlov, 1927; Maren, 2001). The CS is learned as a threat signal (CS+, predicts US). Eventually, the CS+ alone evokes defensive reactions (Fullana et al., 2016; Lonsdorf et al., 2017; Sehlmeyer et al., 2009; Wendt et al., 2017; Heinig et al., 2017; Maren 2001). During extinction training however, the CS+ is repeatedly presented without the US. As a result, the CR decreases gradually due to the formation of new, inhibitory memory traces between the CS+ and no-US. (Graham, 2001; Milad & Quirk, 2012; Heinig et al., 2017). Thus, even after successful extinction training, fear memories can be re-activated (return of fear), e.g., after re-exposure to the US (reinstatement), indicating the co-existence of the initial fear and the newly acquired extinction memories (Bouton, 2004; Hermans et al., 2006; Lonsdorf et al., 2017).

A recent meta-analysis on neural correlates of fear extinction training in healthy adults by Fullana et al. (2018; 31 studies, n=1074 in total) concluded that extinction training activates the brain in a pattern that is reminiscent of fear acquisition (anterior cingulate cortex, the insula, limbic and mid-brain structures; Carvalho et al., 2010; Fanselow 1994; Mobbs et al., 2009; Wendt et al., 2017; Lueken et al., 2013; Sehlmeyer et al., 2011). The authors analyzed studies using uninstructed fear conditioning in differential cue-conditioning paradigms (i.e., a CS+ and a CS− are presented; the CS+ precedes the US) using electric stimulation as US and immediate uninstructed extinction training. Their meta-analysis confirmed the following brain regions to consistently show increased BOLD activation during extinction training in the overall CS+ > CS- comparison: the rostro-dorsal anterior cingulate cortex, medial prefrontal cortex, bilateral anterior insular cortex, frontal operculum, dorsolateral prefrontal cortex, anterior putamen, bilateral ventral pallidum, anterior and medial thalamus, and mid-brain/dorsal pons.

Still missing, though of great importance, is evidence regarding the trajectories of changes in neural activation during the process of extinction training. In general, previous studies (as included in Fullana et al., 2018) focused on neural correlates of conditioned stimuli and their difference contrast (CS+ vs- CS-) during the whole training procedure, but not on changes of neural activation in its course. Due to their methodology of averaging multiple trials for a condition, previous fMRI studies did not prioritize the analysis of activation change trajectories during extinction. However, the neural trajectory may contain important additional information about learning types that could be beneficial for subtyping patients for treatment strategies. Schwarzmeier et al. (2019) found evidence for the importance of neural trajectories in accelerated acquisition of conditioned responses and attenuated extinction recall during delayed extinction training in patients with panic disorder, compared to healthy subjects. Since extinction processes depend on learning competing information challenging well established memories, changes over time are rather subtle and extinction learning is a fragile phenomenon (Bouton, 2002; Giustino and Maren, 2015). Nevertheless, gathering process information is necessary to better understand extinction mechanisms and to disentangle the initial recall of a conditioned fear memory from the progressive formation of the extinction memory (Lonsdorf et al., 2017). In general, these insights would provide a solid basis to inform and optimize exposure-based CBT.

Therefore, we developed an optimized protocol of extinction training closely modeling the conditions and processes of exposure therapy as described in our pilot study (Hollandt et al. 2020). As a delayed extinction paradigm, it models well-consolidated fear memories and includes a reinstatement test, which simulates the relapse of fear symptoms in clinical context. Open questions however remain regarding changes in neural activation across the experimental phases and stability of extinction training effects across repeated measurements in healthy subjects, which is crucial for using the paradigm to investigate treatment associated changes in fear extinction capacities in clinical populations. Furthermore, a given replicability of neural activation patterns on group-level across different study sites would be an important pre-condition for using the paradigm within multi-centric treatment trials.

Goals of the present study were 1) investigating fear-extinction processes during the course of an extinction training at the neural and behavioral level and 2) evaluating the feasibility and replicability of the delayed extinction fMRI-paradigm in a longitudinal-multicenter setting. Thus, this study provides the basis for the investigation of differences in neural fear related mechanisms between patients with anxiety disorders in a clinical context (before and after exposure-based CBT) and healthy subjects. Three main hypotheses about the quality of our paradigm will be investigated:

1. Our newly developed delayed fear extinction paradigm evokes altering BOLD activation in relevant brain regions, especially the cingulate cortex and bilateral insula. Successful extinction learning will be reflected by a decrease of CS+/CS- difference in neural activation and behavioral rating data (US-expectancy, arousal, valence) over the course of the experiment.
2. Reinstatement of conditioned fear elicited by the re-exposure to the US after extinction training will be reflected by a temporary reactivation of CS+/US memory, which will increase the CS+/CS- difference in neural activation and behavioral rating data.
3. We assume that extinction processes can be repeated across time and site setting.

Therefore, a well conducted paradigm will elicit comparable neural processes when applied twice, in spite of familiarity of procedures, context and task in a repeated measurement. This requirement is very important in light of investigations comparing patients before and after exposure-based CBT. Complementarily, we test whether this paradigm can be used in multicentric settings in which data are merged across sites by showing that our paradigm is standardized and robust enough to not produce systematic differences between sites.

## 2. Methods

The current study is part of the national research consortium “Providing Tools for Effective Care and Treatment of Anxiety Disorders” (PROTECT-AD) funded by the German Federal Ministry of Education and Research (BMBF) in the context of the “Research Net for Mental Illnesses” (“Forschungsnetz psychische Erkrankungen”). Using a multicenter randomized controlled trial accompanied by psycho-physiological, neural and (epi-)genetic add-on projects, the consortium aims to test for the critical role of extinction learning during exposure-based CBT (Heinig et al., 2017).

### 2.1 Participants

Quality controlled data-sets of *n* = 100 healthy subjects (46 women; age = 33.1± 10.7 years; 42.4% smokers; higher school education *n* = 75, intermediate *n* = 22, lower *n* = 3) from six sites in Germany with no histories of medical or mental illnesses were included. Information about the quality control process is given in the Supplementary Methods 1.1 and Supplementary Fig. S1. All included subjects had normal or corrected to normal visual acuity and were totally naive to the experiment beforehand. Written informed consent was obtained prior to the participation in the study and the study protocol was approved by the respective local ethics committee according to the Declaration of Helsinki.

### 2.2 Paradigm

The experimental procedure is closely based on a protocol described in detail by Hollandt et al. (2020) tailoring the procedures of exposure therapy. A major experimental optimization is the 24h consolidation phase between fear acquisition and extinction training. Since exposure therapy targets long-lasting and well-consolidated fear memories with a significant delay between fear acquisition and extinction processes in therapy, PROTECT-AD used this delayed extinction paradigm to investigate the mechanisms of extinction training more accurately. Additionally, to make sure that each participant indeed acquired a robust fear response that is supposed to be inhibited by extinction training, PROTECT-AD uses instructed fear acquisition training. In short (see Supplementary Methods 1.2-1.3 for more details), participants underwent a two-day experimental differential fear acquisition and delayed extinction training protocol. Day 1 started with a pre-conditioning phase (both CSs were presented twice without any US presentation). During a subsequent fear acquisition training, one of two neutral facial stimuli (CS+; assignment was counterbalanced between subjects) embedded in either blue or yellow background color co-terminated with an aversive electric US during six of 10 presentations (pseudo-randomized order, 60% reinforcement rate) while the other stimulus (CS−) was never paired with the US during another 10 presentations. Electric stimulation was applied by an electrode attached to the inside of the non-dominant forearm. Immediately before the fear acquisition training, the US intensity was individually calibrated using a standardized shock work-up procedure (Heitland et al., 2013) to find the intensity that was rated as “highly aversive but not painful”. The subjects were instructed about CS+/US contingency (but not the reinforcement rate) prior to the acquisition training. The fear acquisition training took place in an experimental room outside the MRI scanner. Startle blink magnitudes and skin conductance rates (SCR) were assessed on day 1 to check for successful multi-modal acquisition of conditioned fear (Supplementary Methods 1.3 and Supplementary Results 2.1). On day 2, on average 24h after acquisition training, uninstructed extinction training was conducted during fMRI, using the same visual stimuli as CS as on day 1. The US-electrode was attached at the same position as during the previous day and the US intensity was kept the same with an instruction that US-presentation might be possible again. The extinction training started with one re-acquisition trial (one CS+ followed by the US), followed by an extinction training during which both stimuli were presented again 2 x 10 times (first and second extinction phase; Ex1, Ex2) without any US presentation. After that, a reinstatement (RI) of the US comprising the presentation of three USs without any CS presentation during black screen was applied, followed by another 10 presentations for each CS (reinstatement test phase; RIT). During pre-conditioning, fear acquisition, extinction, and reinstatement test, participants were asked to rate US-expectancy, arousal and valence respectively. For the probability estimate of the occurrence of the US during the next trial, a small picture of the upcoming CS informed about the next stimulus to be presented (“Now this picture is following”), accompanied by a request to rate by using the computer mouse (lab, day 1) or keypad (MRI, day 2). On day 1, the US-expectancy ratings preceded every single CS presentation and were conducted on a visual analogue scale (0-100%). US-ratings on day 2 were conducted at six times only in the MRI environment on a 10%-stepped scale: Pre and post re-acquisition, post Ex1, post Ex2, post RI, and post RIT. Ratings of valence and arousal on day 2 were also conducted at six times in the MRI environment on a 10%-stepped scale: Post re-acquisition, pre Ex1, post Ex1, post Ex2, post RI, and post RIT. Since arousal and valence ratings were always given after presentation of the CS (“How aroused/comfortable did you feel during the presentation of the previous picture?”), the Pre Ex1 rating was given after the first of the ten presentations of the CS during Ex1 (for visualization compare Fig. 1). After presentation of the stimuli, requests to rate valence (very bad – very good) and arousal (not agitated at all – very agitated), according to the presented stimulus, were given. Further details about the procedure can be found in the Supplementary Methods 1.2. The whole experimental protocol (t1) was repeated after an interval of an average of 13 weeks (second measurement: t2), using two different visual stimuli as CSs to avoid re-acquisition.

**Figure 1.**
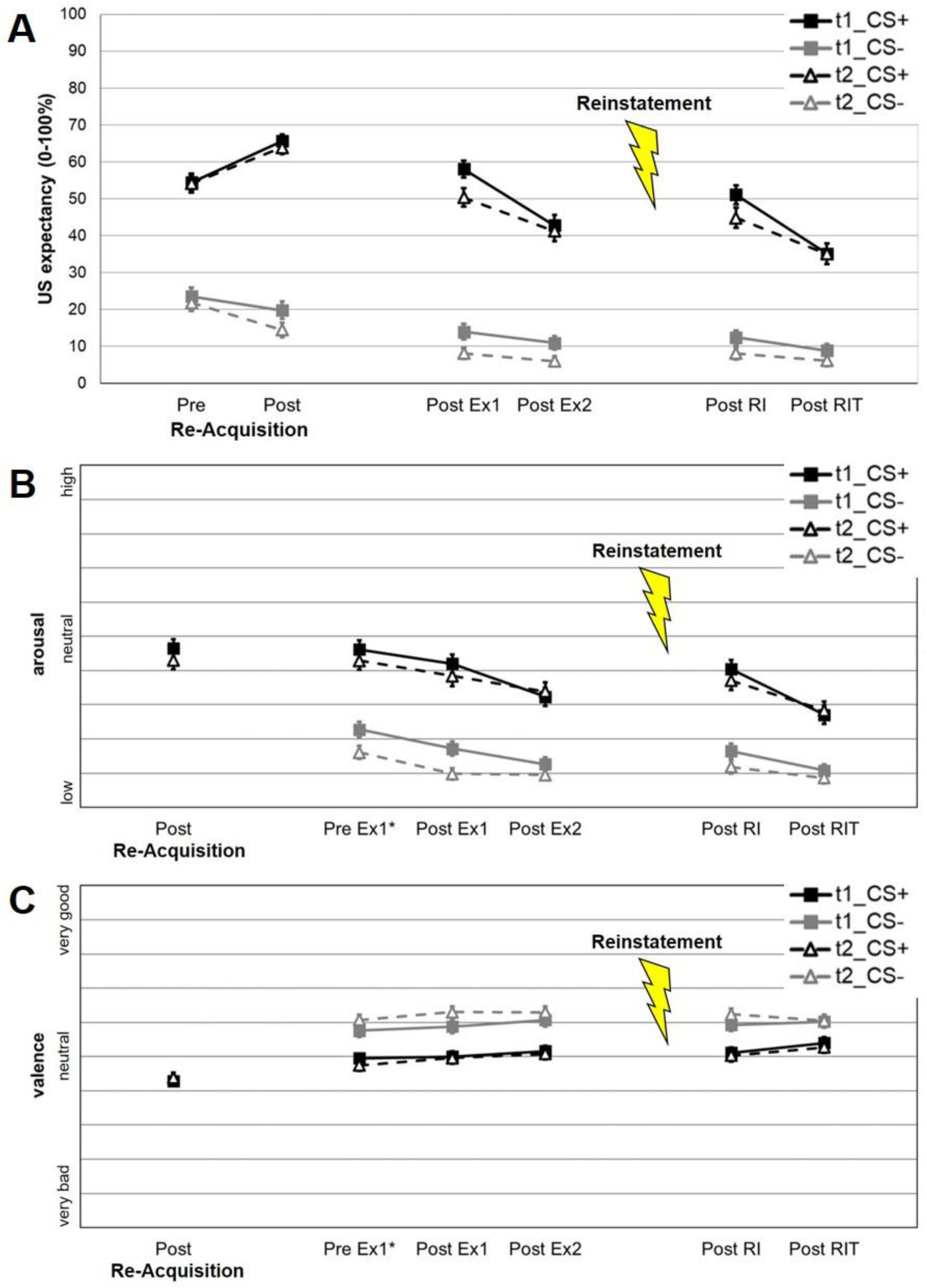
Changes in subjective ratings of CS+ and CS- during extinction training during t1 and t2 for: **A)** US-expectancy **B)** arousal, and **C)** valence. Re-acquisition: One initial pairing of CS+ and US (same intensity as during acquisition training during day 1 confirming contingency. **\***Since arousal and valence ratings were always given after presentation of the CS (“How aroused/comfortable did you feel during the presentation of the previous picture?”), the Pre Ex1 rating was given after the first of the ten presentations of the CS during Ex1.

### 2.3 MRI data acquisition

Comparable to the methods described in Hollandt et al. (2020), time courses of subjects’ brain activity were acquired using 3-Tesla MR scanners (3x Siemens TrioTim, 1x Siemens Verio, 1x Siemens Prisma, 1x Siemens Skyra, Erlangen) equipped with a 12 channel head matrix receive coils. Data quality assurance was guaranteed by harmonized scanner sequences, trained personal, frequent site visits, telephone conferences, and fast online data quality checks with direct feedback to each center (for more information see Supplementary Methods 1.1). Functional images were obtained using a T_2_-weighted gradient-echo echo-planar imaging (EPI) sequence sensitive for the BOLD contrast (TE = 30 ms, TR = 2000 ms, flip angle 90°, matrix size 64 x 64 voxels, voxel size 3.6 x 3.6 x 4.0 mm, slice thickness 4.0 mm, inter-slice gap 0.4 mm, field of view (FOV) = 230 mm, 33 slices, ascending phase encoding direction; due to limitations in technical coordination, a TE = 29 ms had to be used at the Siemens Prisma and at the Siemens Verio 31 slices were recorded). Slices were positioned trans-axially parallel to the intercommissural (AC-PC) plane and tilted 20° to reduce magnetic susceptibility artifacts in prefrontal areas. In total, 590 volumes were collected. In addition, a high-resolution structural image was acquired for each subject using a three-dimensional T_1_-weighted magnetization-prepared rapid gradient-echo (3D MPRAGE) sequence in sagittal plane (TE = 2.26 ms, TR = 1900 ms, inversion time (TI) = 900 ms, flip angle 9°, matrix size 256 x 256 voxels, voxel size 1 x 1 x 1 mm, slice thickness 1.0 mm, FOV = 256 mm, 176 slices; these settings were used in all centers). Total scanning time for the extinction paradigm was 19 min and 4:30 min for the structural scan.

### 2.4 Data analysis

#### 2.4.1 Behavioral data

Behavioral data comprised subjective ratings about US-expectancy, arousal and valence. Statistical data analyses were performed using IBM SPSS v.25 for Linux.

##### Recall of CS+/US-memories

As manipulation, check we used repeated-measures ANOVAs with Stimulus (CS+, CS-) and Time (pre re-acquisition, post re-acquisition) as within-subject factors respectively.

##### Extinction training effects

To investigate effects of extinction training, we used repeated-measures ANOVAs with Stimulus (CS+, CS-) and Time (post re-acquisition/pre Ex1 vs. post Ex1 vs. post Ex2 phase; post RI vs. post RIT) as within-subject factors.

##### Reinstatement effects

For the examination of reinstatement, we used repeated-measures ANOVAs with Stimulus (CS+, CS-) and Time (post Ex2 vs. post RI) as within-subject factors.

##### Effects of measurement point

We assessed the consistency of CS+ > CS-/US-expectancy at pre re-acquisition, post re-acquisition, post Ex1, post Ex2, post RI and post RIT using repeated-measures ANOVAs with Measurement (t1, t2) as within-subjects factor. We tested the consistency of CS+ > CS- arousal and valence at pre Ex1, post Ex1, post Ex2, post RI and post RIT using repeated-measures ANOVAs with Measurement (t1, t2) as within-subjects factor.

##### Effects of site as a covariate

To investigate effects of site we included “site” as a covariate in all ANOVA analyses of CS+/US recall, extinction training effects and reinstatement effects to evaluate, if there were significant interactions.

##### Basic comparisons at sites

For the investigation of effects at each site, we used repeated-measures ANOVAs with Stimulus (CS+, CS-) as within-subject factor for the comparison CS+ > CS- post re-acquisition in US-expectancy separately for each site. Additionally, we used repeated-measures ANOVAs with Stimulus (CS+) as within-subject factor for the comparison post re-acquisition vs. post Ex1 vs. post Ex2. In arousal and valence, we used repeated-measures ANOVAs with Stimulus (CS+, CS-) as within-subject factor for the comparison CS+ > CS- pre Ex1 separately for each site, as well as repeated-measures ANOVAs with Stimulus (CS+) as within-subject factor for the comparison pre Ex1 vs. post Ex1 vs. post Ex2.

##### Exploration of the reliability of behavioral data

To investigate the test-retest-reliability of the behavioral data, we explored Intraclass Correlation Coefficients (ICCs) by means of IBM SPSS Statistics v.25 for Linux. For each domain of interest, a one-way random effects model with single measures (ICC (1,1)) was performed.

##### Recall of CS/US-memories

We analyzed the test-retest-reliability of the CS+/US-expectancy at post re-acquisition and the test-retest-reliability of the CS+ > CS- difference at post re-acquisition. Additionally, the test-retest-reliability of the CS+/arousal and CS+/valence at post re-acquisition and the test-retest-reliability of the CS+ > CS- difference in arousal and valence at pre Ex1 was studied.

##### Extinction training effects

We analyzed the test-retest-reliability of the CS+ extinction training effect in US-expectancy from post re-acquisition over post Ex1 to post Ex2 and the test-retest-reliability of extinction training effect of the CS+ > CS- difference from post re-acquisition over post Ex1 to post Ex2. We analyzed the test-retest-reliability of the CS+ extinction training effect in arousal and valence from post re-acquisition over pre Ex1 and post Ex1 to post Ex2 and the test-retest-reliability of extinction training effect in arousal and valence of the CS+ > CS- difference from pre Ex1 over post Ex1 to post Ex2.

##### Reinstatement

We analyzed the test-retest-reliability of the CS+ > CS- difference post Ex2 and post RI separately, as well as the difference post Ex2 vs. post RI in the CS+ > CS- difference for US-expectancy, arousal and valence.

#### 2.4.2 fMRI data

Functional MRI data were preprocessed and analyzed using the SPM12b software package (Statistical Parametric Mapping, Wellcome Trust Center for Neuroimaging, London, UK; http://www.fil.ion.ucl.ac.uk), based on Matlab R2009b (version 7.9.0; MathWorks). After discarding the first five volumes to minimize T1-saturation effects, all images were high-pass filtered (cut-off period 128 s), co-registered to the anatomical T1 images, spatially realigned and normalized into the MNI space using the MNI template (resulting voxel size 2 x 2 x 2 mm^3^). Spatial smoothing followed an iterative approach recommended for multi-center studies (Friedman et al., 2006) with a target kernel of 8 mm isotropic Gaussian filter. As iterative smoothing procedures take differences in intrinsic smoothness (Friston et al., 1994) of the data into account, it might reduce variance induced by smoothness even in context of a single center study.

Statistical whole-brain analysis was performed in a two-level, mixed-effects procedure. Voxel-wise BOLD activity in our event-related design was modeled by means of a single subject first-level General Linear Model (GLM) comprising the onsets of the US, rating phases and CS+/CS- for each experimental phase (10 trials for each stimulus type in each experimental phase) respectively, resulting in eight experimental regressors, convolved with the HRF. The six realignment parameters were additionally included as nuisance covariates to account for residual motion artifacts. The hemodynamic response was modeled by the canonical hemodynamic response function (HRF). Parameter estimate (*β*-) images for the HRF were calculated for each condition and each subject. Parameter estimates for the twelve relevant conditions (CS+ and CS- for each block [Ex1, Ex2, RIT] respectively for measurement point t1 and t2) were then entered into a within-subject flexible factorial analysis. Site was added as a covariate. Data from both measurement points (t1 + t2) and all sites were integrated in one 2^nd^-level-analysis to enable investigation of time-dependent and time-independent effects. For further site-specific analyses, additional 2^nd^-level-analyses for each individual site were performed.

A Monte Carlo simulation (Slotnik, 2017) at threshold *p* < .005 (uncorr.) with a minimum cluster size of 175 contiguous voxels was used to correct for multiple comparisons at *p* < .05 for all contrasts. Additional family-wise error (FWE) correction for each peak and cluster as calculated by SPM will be provided in all tables.

##### Contrasts of interest

###### Extinction training effects

Extinction training effects regarding the reduction of the conditioned response (CS+ > CS- difference) from Ex1 to Ex2 (two-way interaction of CS-type [CS+ > CS-] X Block [Ex1 > Ex2]) and correspondingly, extinction training effects regarding the changes in BOLD suppression on CS+ (CS- > CS+ difference) from Ex1 to Ex2 (two-way interaction of CS-type [CS+ < CS-] X Block [Ex1 > Ex2]) were investigated each by a *T*-contrast. Both measurement points were combined to increase statistical power.

###### Effects of measurement point

Potential differential effects of the measurement point on extinction training effects were investigated using three-way interaction analyses (*F*- contrasts): Measurement (t1, t2) X CS-type (CS+, CS-) X Block (Ex1, Ex2) and Measurement (t1, t2) X CS-type (CS+, CS-) X Block (Ex2, RIT). Additional conjunction analyses (minimum t statistic, conjunction null) on the consistency of the t1 and t2 fMRI data were applied in the respective contrasts CS+ > CS- as well as CS- > CS+ during Ex1.

###### Effects of site as a covariate

*F*-contrast of interest regarding the overall effects of site was operationalized by exploring the combined effects of regressors put on the “site” covariates against implicit baseline.

###### Basic comparisons at site

*T*-contrasts of interest for each individual site were (CS+ > CS-) and (CS- > CS+) during Ex1, respectively. Both measurement points were combined to increase statistical power.

###### Reinstatement effect in fMRI data

Effects regarding the activation changes between Ex2 and RIT were calculated by *T*-contrasts in both directions: ([CS+ > CS-] X [Ex2 vs. RIT]) and ([CS- > CS+] X [Ex2 vs. RIT]). Both measurement points were combined to increase statistical power.

To explore specific effects of the reinstatement between the end of Ex2 and the beginning of RIT, we modeled this part of the experiment in a separate analysis. Knowing that reinstatement effects in healthy subjects are small and of short duration (Haaker et al., 2014), a single subject first-level GLM was conducted comprising the onsets of the US, rating phases and respectively only five CS+/CS- stimuli of the late half of Ex2 and the early half of RIT, resulting in six experimental regressors plus the six realignment parameters. Parameter estimates for the eight relevant conditions (CS+ and CS- for each half block of the Ex2 and RIT respectively for t1 and t2) were then entered as within-subject conditions in a flexible factorial analysis. Site was added as a covariate. *F*-Contrast of interest was the interaction of CS-type X pre/post reinstatement. Again, both measurement points were combined to increase statistical power.

###### Exploration of the reliability of fMRI data

To get a general idea of the test-retest-reliability of the fMRI data, we explored Intraclass Correlation Coefficients (ICCs) by means of IBM SPSS Statistics v.25 for Linux on extracted eigenvariates from the 2nd-level. To avoid confounds with measurement point as within-subject factor, we conducted two separate analyses including only t1-data and only t2-data without any covariates, respectively. We built the contrast CS+ > CS- during Ex1. From the CS+ > CS- contrast in Ex1, we then extracted eigenvariates from clusters activated in this contrast in those regions that also reflected relevant CS+ > CS- difference reduction in the previous analysis of extinction training effects (see 3.3.2 “Exploration of the reliability of fMRI data” for details on these ROIs). Eigenvariates were extracted over all conditions (CS+ and CS- for each block [Ex1, Ex2, RIT]), respectively from the t1-data and t2-data 2nd-level analysis. […]”

To correct for effects of site, regression analyses for all extracted values were performed, predicting eigenvariates by site. Standardized predicted values and residuals were saved. All ICC calculations were based on the residual values not explained by site. For each region and each domain of interest a one-way random effects model with single measures (ICC (1,1)) was performed.

###### Recall of CS/US-memories

We analyzed the test-retest-reliability of the CS+ during Ex1 and the test-retest-reliability of the CS+ > CS- difference during Ex1.

###### Extinction training effects

We analyzed the test-retest-reliability of the CS+ extinction training effect (Ex1 > Ex2) and the test-retest-reliability of extinction training effect of the CS+ > CS- difference (Ex1 > Ex2).

###### Reinstatement effect

We analyzed the test-retest-reliability of the CS+ > CS- difference each separately for pre and post reinstatement as well as the test-retest-reliability of the interaction pre vs. post reinstatement in the CS+ > CS- difference.

###### Exploration of correlations between fMRI and behavioral data

To explore the association between neural activation changes and changes in subjective ratings, we calculated bivariate Pearson correlations in IBM SPSS Statistics v.25 for Linux between the extracted eigenvariates from clusters with significant activation and the rating data.

## 3. Results

Fig. 1 provides a visualization of US-expectancy (A), arousal (B) and valence (C) ratings during extinction training (day 2). To provide evidence for successful fear acquisition on day 1 we also report results from subjective ratings, skin conductance response (SCR) and startle reaction from fear acquisition training (Supplementary Results 2.1 and Supplementary Fig. S2), and analyzed the recall of the conditioned response and the effect of re-acquisition before extinction training (Supplementary Results 2.2-2.3). As additional manipulation check in fMRI data, we also analyzed the main effects (CS+ > CS-) and (CS- > CS+) during Ex1 at t1 and t2 respectively (Supplementary Table S1).

### 3.1 Extinction training effects

#### 3.1.1 Behavioral data

##### US-expectancy

A decrease in CS+ > CS- differentiation was reflected in a Time X Stimulus interaction from post re-acquisition to post Ex2 ((re-acquisition > post Ex1 > post Ex2) x (CS+ > CS-); at t1: *F*(2,98) = 15.68, *p* < .001, *η_p_^2^* = 0.14; at t2: *F*(2,98) = 19.06, *p* < .001, *η_p_^2^* = 0.16). From post re-acquisition to post Ex2, the CS+/US-expectancy decreased ((re-acquisition > post Ex1 > post Ex2) x CS+; at t1: *F*(2,98) = 43.36, *p* < .001, *η_p_^2^* = 0.31; at t2: *F*(2,98) = 48.62, *p* < .001, *η_p_^2^* = 0.33; Fig.1A).

##### Arousal

The CS+ related arousal decreased from pre Ex1 to post Ex2 ((pre Ex1 > post Ex1 > post Ex2) x CS+; at t1: *F*(2,98) = 26.45, *p* < .001, *η_p_^2^* = 0.21; at t2: *F*(2,98) = 9.36, *p <* .001, *η_p_^2^* = 0.09; Fig.1B).

##### Valence

The CS+ related negative valence reduced only at t2 from pre Ex1 to post Ex2 ((pre Ex1 > post Ex1> post Ex2) x CS+ at t2: F(2,98) = 4.26, p = .02, *η_p_^2^* = 0.04), while there was no significant reduction at t1 ((pre Ex1 > post Ex1 > post Ex2) x CS+ at t1: F(2,98) = 1.20, p = .30, *η_p_^2^* = 0.01). At the very end of the experiment, valence ratings were still significantly lower for CS+ compared to CS- (post RIT x (CS+ > CS-); at t1: *F*(1,98) = 14.77, *p* < .001, *η_p_^2^* = 0.13; at t2: *F*(1,98) = 22.48, *p <* .001, *η_p_^2^* = 0.19; Fig.1C).

#### 3.1.2 fMRI

Across both measurement points (t1 + t2), we found extinction training effects from first to second extinction phase (Ex1 vs. Ex2) regarding the decreasing CS+ > CS- difference (mainly driven by an activation decrease corresponding to the CS+) in the insular cortex and parietal operculum with supramarginal gyrus as well as in the middle cingulate cortex ((Ex1 > Ex2) x (CS+ > CS-), Table 1A, Fig. 2).

**Table 1.** Extinction effects from the first to the second extinction phase and from the second extinction phase to the post-reinstatement phase, respectively, across both measurement points. All contrasts were assessed at *p* < .005 uncorrected with a cluster threshold of *k* = 175.

| Contrast/Region | hemi-<br>sphere | no.<br>voxels | MNI<br>coordinates<br>peak |  |  |  | P-value FWE-corrected |  |
| --- | --- | --- | --- | --- | --- | --- | --- | --- |
|  |  |  | x | y | z | t | peak | cluster |
| **A) t1+t2: Ex1 (CS+ > CS-) vs. Ex2 (CS+ > CS-)** |  |  |  |  |  |  |  |  |
| Postcentral gyrus + supramarginal gyrus + parietal operculum | L | 647 | -64 | -22 | 26 | 4.93 | .014 | .009 |
| Parietal operculum + supramarginal gyrus | R | 901 | 44 | -24 | 26 | 4.31 | .170 | .001 |
| Central operculum + anterior insula | L | 523 | -46 | 6 | 10 | 4.03 | .404 | .025 |
| Middle cingulate gyrus | R | 1838 | 4 | 12 | 32 | 3.97 | .472 | <.001 |
| Postcentral gyrus | R | 351 | 26 | -38 | 70 | 3.89 | .566 | .118 |
| Cerebellum | L | 424 | -2 | -60 | -24 | 3.88 | .574 | .060 |
| Central operculum + anterior insula | R | 228 | 48 | 2 | 4 | 3.70 | .784 | .377 |
| **B) t1+t2: Ex1 (CS- > CS+) vs. Ex2 (CS+ > CS-)** |  |  |  |  |  |  |  |  |
| Precuneus + posterior cingulate cortex | L | 451 | -6 | -52 | 32 | 3.79 | .683 | .047 |
| **C) t1+t2: Ex2 (CS+ > CS-) vs. RIT (CS+ > CS-)** |  |  |  |  |  |  |  |  |
| Anterior cingulate gyrus | R | 507 | 8 | 36 | 18 | 4.67 | .044 | .029 |
| Anterior insula | R | 312 | 30 | 22 | -10 | 4.61 | .056 | .171 |
| **D) t1+t2: Ex2 (CS- > CS+) vs. RIT (CS+ > CS-)** |  |  |  |  |  |  |  |  |
| Pre- + postcentral gyrus | L | 14566 | 12 | -36 | 64 | 6.41 | <.001 | <.001 |
| Occipital gyrus | L | 4388 | 22 | -76 | 34 | 4.77 | .028 | <.001 |
| Cerebellum | L | 175 | -12 | -60 | -36 | 3.80 | .681 | .593 |
| Hippocampus + parahippocampal gyrus + fusiform gyrus | L | 283 | -26 | -32 | -12 | 3.65 | .834 | .226 |
| Inferior + middle frontal gyrus | R | 177 | 48 | 32 | 10 | 3.58 | .893 | .584 |

**Abbreviations:** CS+: conditioned stimulus that is followed by the unconditioned stimulus (US) with a reinforcement rate of 60% (only unpaired CS+ were included); CS-: conditioned stimulus that is never followed by an US; L: left; R: right; no. voxel: number of voxels per cluster; x, y, z: MNI coordinates.

**Figure 2.**
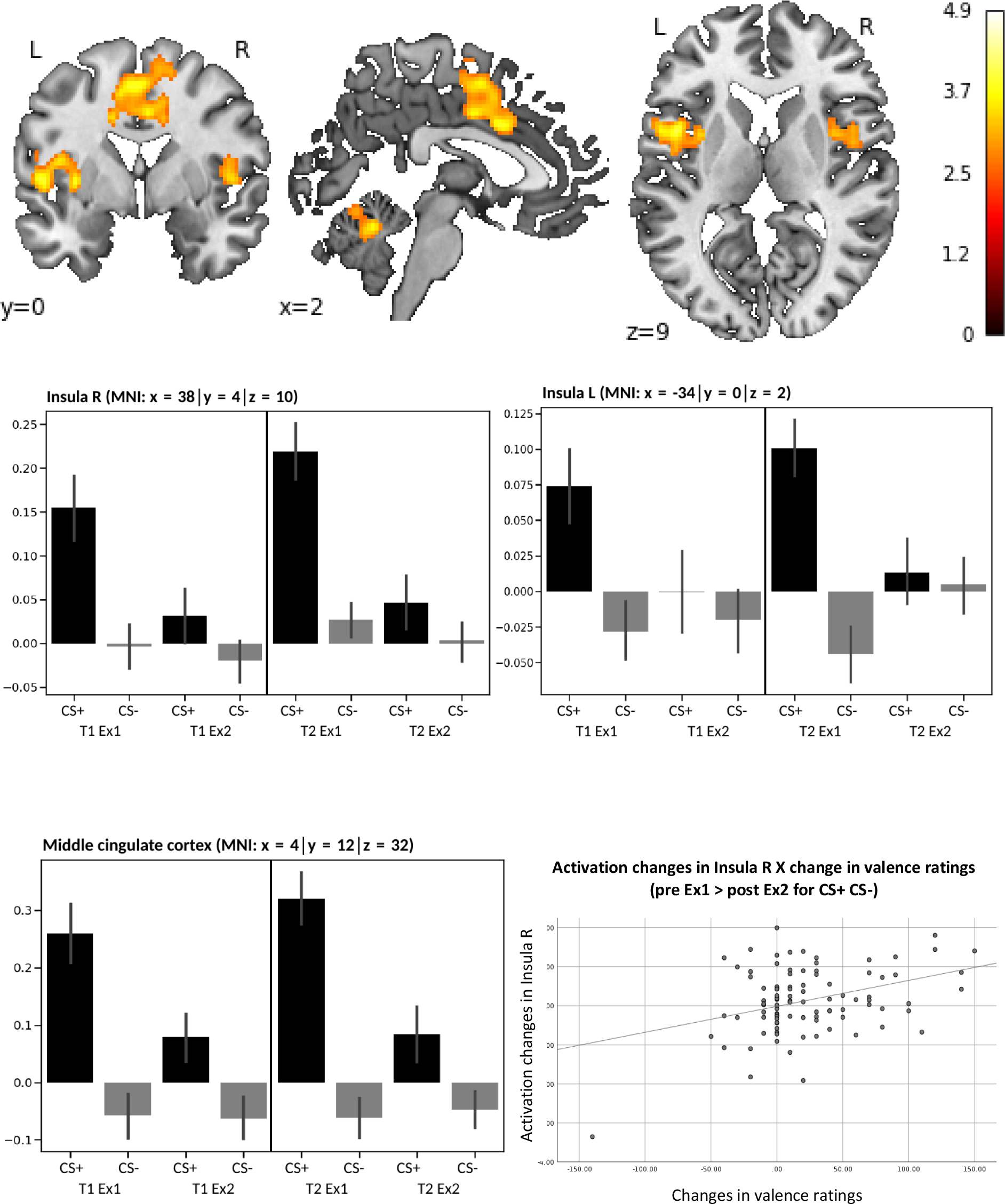
Activation difference CS+ > CS- form first (Ex1) to second (Ex2) extinction phase. Correlation of activation change in the right insula (pre Ex1 > post Ex2 for CS+ CS-) with the change in valence ratings (pre Ex1 > post Ex2 for CS+ CS-). All contrasts were calculated across both measurement points and assessed at *p* < .005 uncorrected with a cluster threshold of *k* = 175 (see Table 1).

Regarding the CS- > CS+ difference (mainly driven by an activation increase corresponding to the CS+), we found activation in a cluster of the left precuneus extending to the posterior cingulate cortex (PCC) ((Ex1 > Ex2) x (CS- > CS+), Table 1B, Fig. 3).

**Figure 3.**
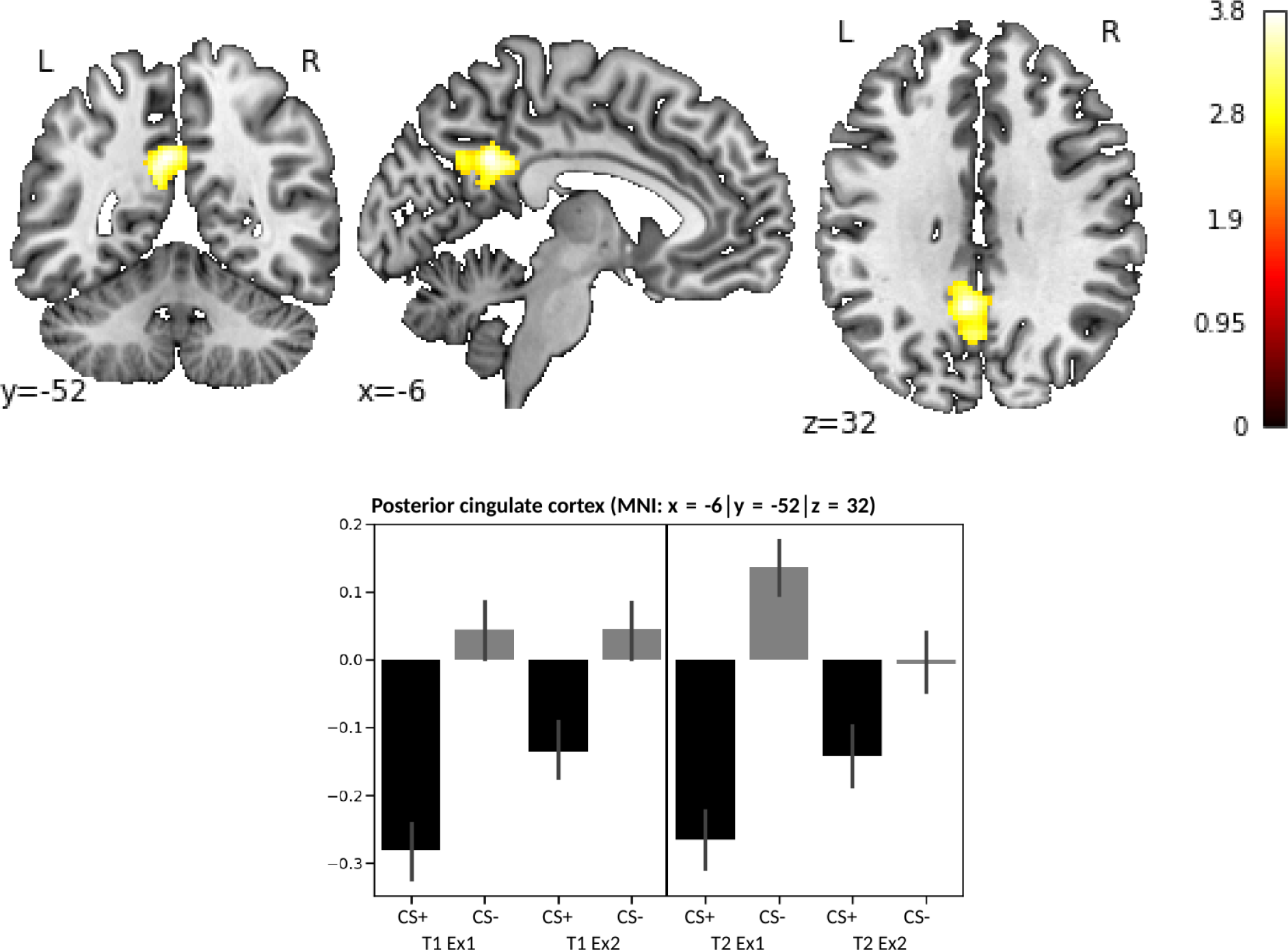
Activation difference CS- > CS+ form first (Ex1) to second (Ex2) extinction phase. All contrasts were calculated across both measurement points and assessed at *p* < .005 uncorrected with a cluster threshold of *k* = 175 (see Table 1).

Activation change (Ex1 > Ex2 for CS+ > CS-) in the bilateral insula and middle cingulate cortex was correlated with change in valence ratings (pre Ex1 > post Ex1 > post Ex2 for CS+ > CS-): right insula *r* = .36, *p* < .001 (Fig. 2); left insula *r* = .32, *p* = .001; cingulate cortex *r* = .24, *p =* .018. No correlation was found with changes in US-expectancy and arousal ratings.

### 3.2 Reinstatement effects

#### 3.2.1 Behavioral data

##### US-expectancy

A reinstatement effect in CS+/CS- differentiation was observed only during t1 in a Time X Stimulus interaction from post Ex2 to post RI ((post Ex2 > post RI) x (CS+ > CS-), at t1: *F*(1,98) = 10.69, *p* = .001, *η_p_^2^* = 0.10; at t2: *F*(1,98) = .78, *p* = .38, *η_p_^2^* = 0.01). The reinstatement led to an increased CS+/US-expectancy ((post Ex2 > post RI) x CS+ at t1: *F*(1,98) = 18.49, *p* < .001, *η_p_^2^* = 0.16; at t2: *F*(1,98) = 6.71, *p* = .011, *η_p_^2^* = 0.06) which then decreased again after RIT ((post RI > post RIT) x CS+ at, t1: *F*(1,98)=46.64, *p*<.001, *η_p_^2^* =.32; at t2: *F*(1,98) = 39.81, *p <* .001, *η_p_^2^* = .29). At the end of the experiment, the CS+/US-expectancy as well as the CS-/US-expectancy were still higher than 0 and a CS+ > CS- differentiation regarding US-expectancy was still present (post RIT x (CS+ > CS-), at t1: *F*(1,98) = 69.77, *p* < .001, *η_p_^2^* = 0.42; at t2: *F*(1,98) = 93.74, *p <* .001, *η_p_^2^* = 0.49; Fig.1A).

##### Arousal

After the reinstatement, the CS+ related arousal increased again ((post Ex2 > post RI) x CS+, at t1: *F*(1,98) = 19.84, *p* < .001, *η_p_^2^* = 0.17; at t2: *F*(1,98) = 4.06, *p* = .05, *η_p_^2^* = 0.04) and decreased during RIT ((post RI > post RIT) x CS+, at t1: *F*(1,98) = 48.45, *p* < .001, *η_p_^2^* = 0.33; at t2: *F*(1,98) = 27.60, *p <* .001, *η_p_^2^* = 0.22). At the very end of the experiment arousal was still rated significantly higher for CS+ compared to CS- (post RIT x (CS+ > CS-), at t1: *F*(1,98) = 44.61, *p* < .001, *η_p_^2^* = 09.31; at t2: *F*(1,98) = 60.75, *p <* .001, *η_p_^2^* = 0.38; Fig.1B).

##### Valence

There were no significant changes in valence ratings.

#### 3.2.2 fMRI

Between the second extinction phase and the reinstatement test phase, effects in a decrease in CS+ > CS- difference were found in the anterior insular and anterior cingulate cortex ((Ex2 > RIT) > (CS+ > CS-), Table 1C, Fig. 4).

**Figure 4.**
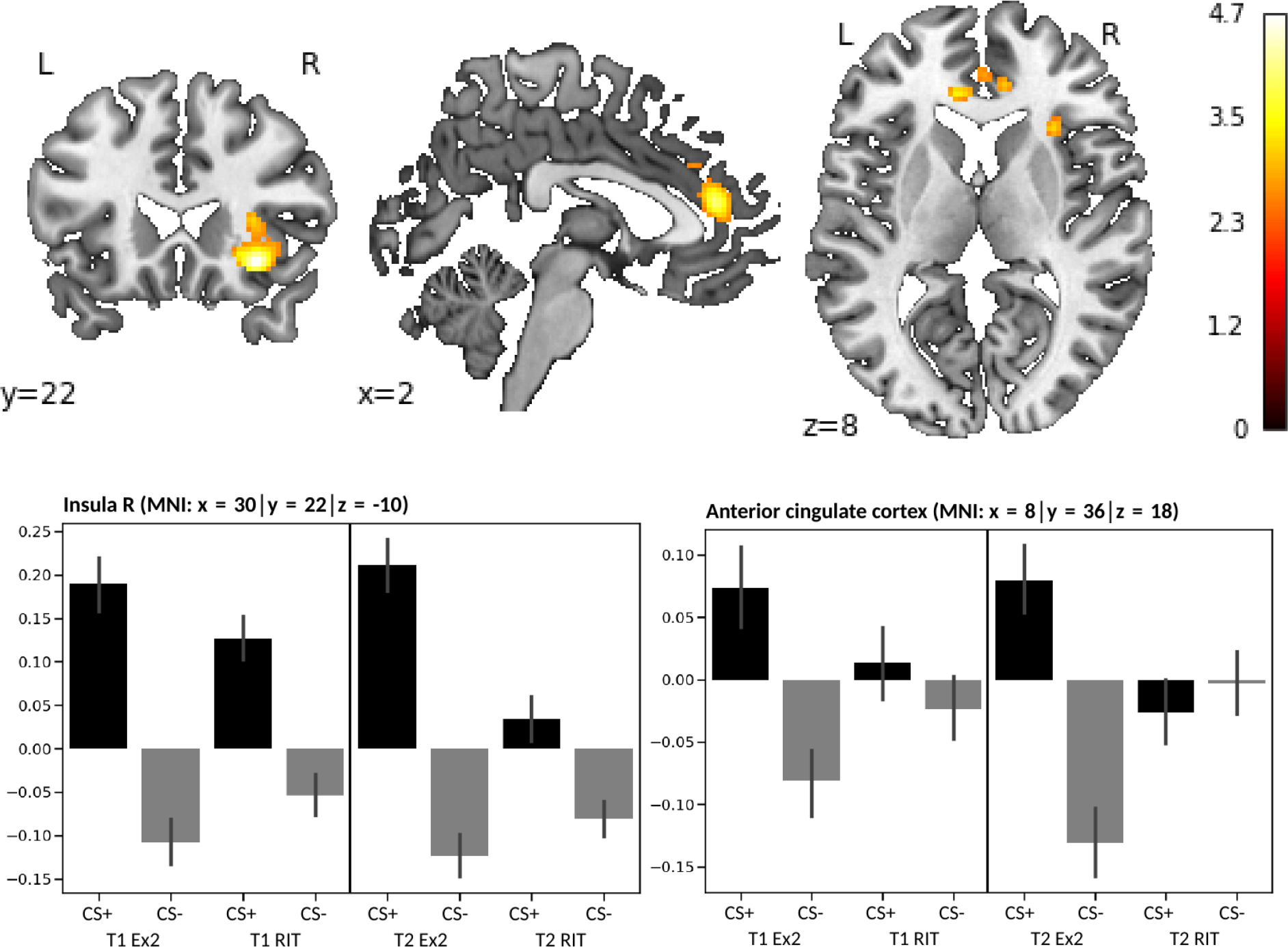
Activation difference CS+ > CS- form second extinction phase (Ex2) to post-reinstatement phase. All contrasts were calculated across both measurement points and assessed at *p* < .005 uncorrected with a cluster threshold of *k* = 175 (see Table 1).

An increase in activation towards CS+ was found in the CS- > CS+ difference contrast predominately in the bilateral pre- and postcentral gyrus, occipital areas and cerebellum, as well as in the left hippocampus ((Ex2 > RIT) > (CS- > CS+), Table 1D, Fig. 5)”.

**Figure 5.**
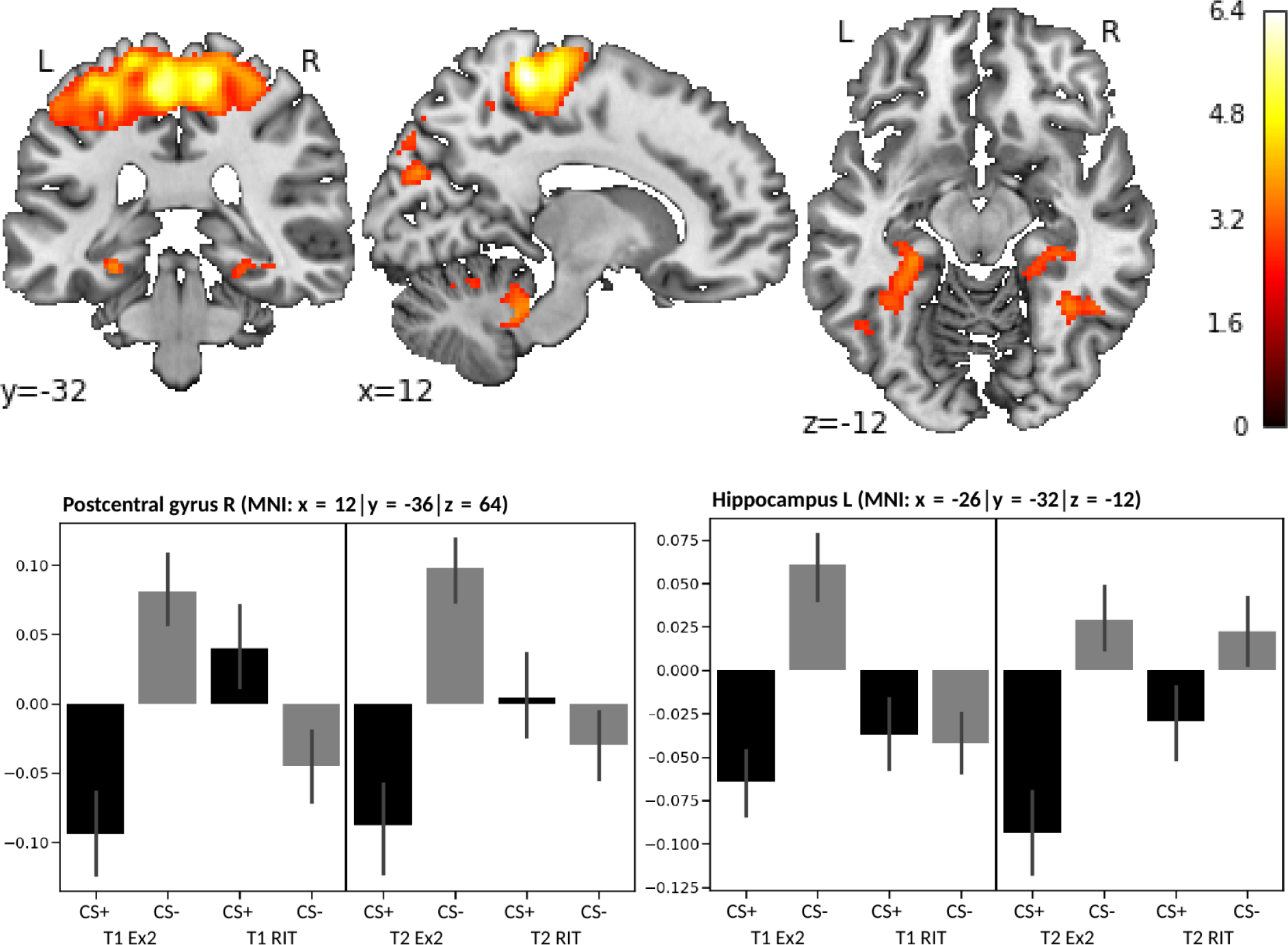
Activation difference CS- > CS+ from second extinction phase (Ex2) to post-reinstatement phase. All contrasts were calculated across both measurement points and assessed at *p* < .005 uncorrected with a cluster threshold of *k* = 175 (see Table 1).

In the additional analysis with only the late half of Ex2 and early half of RIT (5-trials each), no sig. results on our corrected threshold were found. However, anticipating small effects, we decided to explore this contrast at threshold *p* < .005 (uncorr.) with no minimum cluster size. On the uncorrected level however, we found the following: Across both measurement points (t1 + t2) we found a reinstatement effect in an interaction of CS-type X pre/post reinstatement in a cluster of the ACC ((pre RI vs post RI) x (CS+ vs. CS-), voxel size: 109; x = 0; y = 40; z = 10; *F* = 16.60; FWE-corrected *p_peak level_* = .605; *p_uncorrected_* < .001, threshold *p* < .005). Before the reinstatement, BOLD-activation in this cluster was higher during CS- presentation than during CS+. After the reinstatement, this differentiation showed the opposite pattern (Supplementary Fig. S3).

Activation change (pre reinstatement vs. post reinstatement for CS+ vs. CS-) in the ACC was correlated with change in US-expectancy ratings (pre reinstatement vs. post reinstatement for CS+ > CS-): *r* = .25, *p* = .013. No correlation was found with changes in arousal and valence ratings.

### 3.3 Stability of paradigm

#### 3.3.1 Stability of behavioral data

##### Effects of time

The US-expectancy, arousal and valence rating data showed the same pattern of CS+ > CS- differences (Supplementary Table S2) at both measurement points (t1, t2).

##### Effects of site as a covariate

In neither analysis of CS+/US recall (see Supplementary Results) nor extinction training effects (see 3.1.1) and reinstatement effects (see 3.2.1) regarding US-expectancy, arousal and valence, was there any significant interaction with the “site” covariate to be found.

##### Basic comparisons at sites

###### CS+ > CS-

At all sites respectively and across both measurement points (t1 + t2), a significant CS+ > CS- difference after re-acquisition in US-expectancy and at pre Ex1 in arousal and valence was found (Supplementary Table S3).

###### CS+ extinction

At five of six sites a significant decrease in CS+/US-expectancy from the re-acquisition over post Ex1 to post Ex2 was found across both measurement points (t1 + t2). At three of six sites, a significant decrease in CS+/arousal from the re-acquisition over pre Ex1 and post Ex1 to post Ex2 was found across both measurement points (t1 + t2). At none of the six sites, a significant decrease in CS+/valence was found (Supplementary Table S3).

##### Exploration of the reliability of behavioral data

###### CS+/US-expectancy

The test-retest-analysis revealed fair reliability of both the CS+/US-expectancy at post re-acquisition was (*ICC* = .51, *F*(98,99) = 3.11, *p* < .001) and the CS+ > CS- difference in CS+/US-expectancy at post re-acquisition (*ICC* = .49, *F*(98,99) = 2.88, *p* < .001).

###### Arousal

We found fair reliability for the CS+/arousal at post re-acquisition (*ICC* = .53, *F*(98,99) = 3.22, *p* < .001) as well as for the CS+ > CS- difference in arousal at pre Ex1 (*ICC* = .42, *F*(98,99) = 2.42, *p* < .001).

###### Valence

For CS+/valence at post re-acquisition, the reliability was fair (*ICC* = .49, *F*(98,99) = 2.92, *p* < .001). There was no significant test-retest-reliability of the CS+ > CS- difference in valence at pre Ex1 (*ICC* = .02, *F*(98,99) = 1.04, *p* = .42).

##### Extinction training effects

###### CS+/US-expectancy

For the CS+ extinction training effects in CS+/US-expectancy effect from post re-acquisition over post Ex1 to post Ex2 (*ICC* = .66, *F*(98,99) = 4.91, *p* < .001), as well as for the extinction training effect of the CS+ > CS- difference from post re-acquisition over post Ex1 to post Ex2, the test-retest-reliability was good (*ICC* = .64, *F*(98,99) = 4.62, *p* < .001).

###### Arousal

For the CS+ extinction training effects in arousal from post re-acquisition over pre Ex1 and post Ex1 to post Ex2, the test-retest-reliability was good (*ICC* = .63, *F*(98,99) = 4.37, *p* < .001). For the extinction training effect of the CS+ > CS- difference from pre Ex1 over post Ex1 to post Ex2 test-retest-reliability was fair (*ICC* = .43, *F*(98,99) = 2.49, *p* < .001).

###### Valence

The test-retest-reliability of the CS+ extinction training effects in valence effect from post re-acquisition over pre Ex1 and post Ex1 to post Ex2, was fair (*ICC* = .56, *F*(98,99) = 3.55, *p* < .001), but the test-retest-reliability of extinction training effect of the CS+ > CS- difference from pre Ex1 over post Ex1 to post Ex2 was low (*ICC* = .25, *F*(98,99) = 1.66, *p* = .006).

##### Reinstatement

###### US-expectancy

The test-retest-reliability of the CS+ > CS- difference pre reinstatement was good (*ICC* = .67, *F*(98,99) = 5.14, *p* < .001) and post reinstatement was fair (*ICC* = .52, *F*(98,99) = 3.19, *p* < .001). For the difference pre vs. post reinstatement in the CS+ > CS- difference, the reliability was low (*ICC* = .22, *F*(98,99) = 1.58, *p* = .012).

###### Arousal

The test-retest-reliability of the CS+ > CS- difference pre reinstatement (*ICC* = .53, *F*(98,99) = 3.24, *p* < .001) and post reinstatement was fair (*ICC* = .55, *F*(98,99) = 3.47, *p* < .001). For the difference pre vs. post reinstatement in the CS+ > CS- difference, the reliability was low (*ICC* = .19, *F*(98,99) = 1.46, *p* = .031).

###### Valence

For the CS+ > CS- difference pre reinstatement (*ICC* = .39, *F*(98,99) = 2.28, *p* < .001), as well as post reinstatement, the test-retest-reliability was low (*ICC* = .34, *F*(98,99) = 2.03, *p* < .001). There was no significant ICC for the difference pre vs. post reinstatement in the CS+ > CS- difference (*ICC* = -.03, *F*(98,99) = .94, *p* = .62).

#### 3.3.2 Stability of fMRI data

##### Effects of time

The three-way interactions of CS type (CS+, CS-) X time (Ex1, Ex2) X measurement point (t1, t2) and CS type X time (Ex2, RIT) X measurement point revealed no significant effects (threshold p < .005; k = 175), suggesting similar BOLD activation patterns due to extinction training and reinstatement between t1 and t2.

We further evaluated the consistency of the t1 and t2 fMRI data by analyzing the conjunction (minimum t-statistics, conjunction null) of the respective differential effects CS+ > CS- during Ex1 showing high consistency in insular, cingulate and supramarginal activation and CS- > CS+ during Ex1 showing high consistency in the precuneus with posterior cingulate cortex, the medial frontal gyrus with gyrus rectus including the area of the ventromedial prefrontal cortex, angular gyrus and hippocampal areas (Supplementary Table S4, Supplementary Fig. S4 + S5).

##### Effects of site as a covariate

Differences in neural activation due to the different sites were found predominately in occipital areas as well as in the cerebellum, thalamus and basal ganglia (Fig. 6). Detailed information can be found in Table 2. Since there seemed to be some overlap in activity with the CS- > CS+ extinction effects from Ex2 to RIT, we explored the potential influence of site effects on findings from Ex2 to reinstatement in a conjunction analysis (minimum t statistic, conjunction null). We found an overlap of site effects with the (CS- > CS+) x (Ex2 > RIT) contrast related activity in a cluster of the occipital cortex (k = 216, MNI-coordinates x = 26; y = −80; z = 28, t = 3.79, p_peak level_ = .688 FWE-corrected, assessed at p<.005 uncorrected and a cluster threshold of k=175).

**Table 2.** Differences in neural activation due to the different centers. Effects across the whole experiment. All contrasts were assessed at *p* < .005 uncorrected and a cluster threshold of *k* = 175.

| Region | hemi-<br>sphere | no.<br>voxels | MNI coordinates<br>peak |  |  |  | P-value FWE-corrected |  |
| --- | --- | --- | --- | --- | --- | --- | --- | --- |
|  |  |  | x | y | z | F | peak | cluster |
| Occipital gyrus + occipital pole | R | 3191 | 28 | -94 | 6 | 106.58 | <.001 | <.001 |
| Lingual gyrus + calcarine cortex | R | 1541 | 6 | -78 | 0 | 87.16 | <.001 | <.001 |
| Superior occipital + Cuneus + superior occipital gyrus + occipital pole | L | 466 | -16 | -94 | 20 | 71.24 | <.001 | .015 |
| Cerebellum | L | 309 | -16 | -76 | -22 | 37.55 | <.001 | .090 |
| Cerebellum + occipital fusiform gyrus | R | 755 | 38 | -70 | -24 | 3.02 | .002 | .001 |
| Fusiform gyrus | L | 265 | -34 | -56 | -8 | 27.05 | .008 | .153 |
| Caudate | R | 297 | 20 | 16 | 12 | 15.15 | .809 | .104 |

**Abbreviations:** CS+: conditioned stimulus that is followed by the unconditioned stimulus (US) with a reinforcement rate of 60% (only unpaired CS+ were included); CS-: conditioned stimulus that is never followed by an US; L: left; R: right; no. voxel: number of voxels per cluster; x, y, z: MNI coordinates.

**Figure 6.**
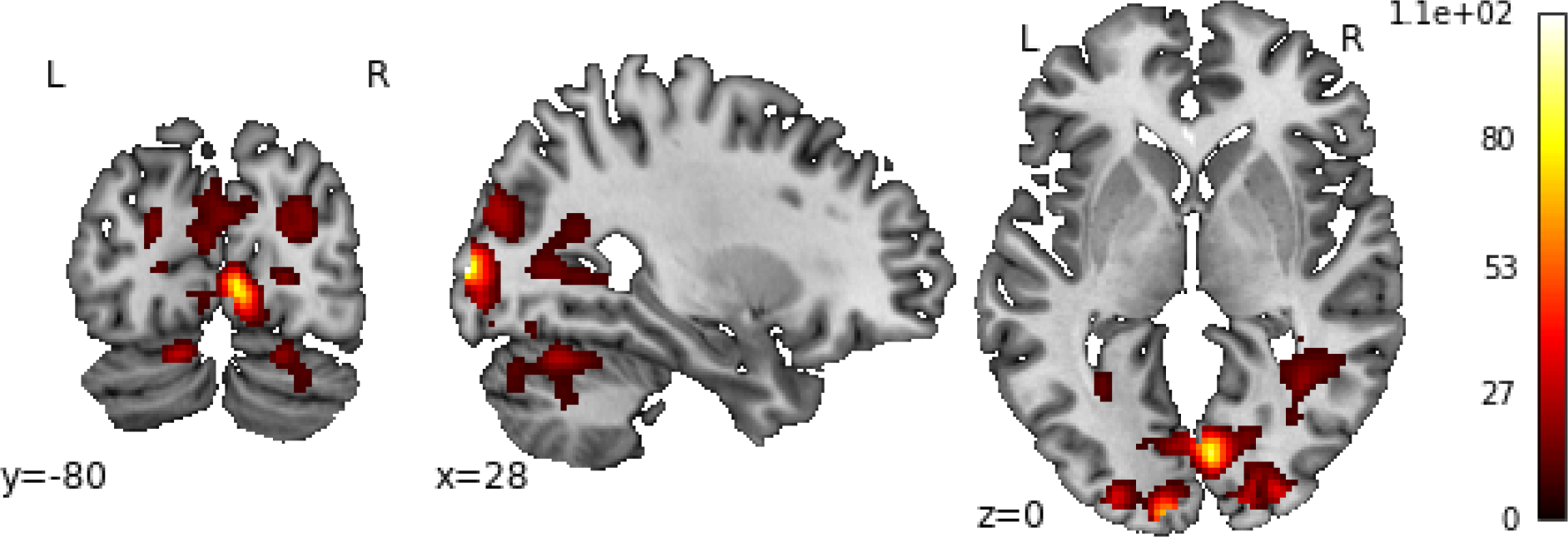
Differences in neural activation due to the different centers. Effects across the whole experiment. All contrasts were assessed at *p* < .005 uncorrected with a cluster threshold of *k* = 175 (see Table 2).

##### Basic comparisons at sites

Detailed information can be found in the Supplementary Tables S5-10. Across both measurement points (t1 + t2), activation differences between the learned threat and safety signal (CS+ > CS- and CS- > CS+) during the first extinction phase were assessed for each site individually. At every site, CS+ > CS- differences were found in the anterior insular cortex, middle cingulate cortex and supramarginal gyrus, whereas CS- > CS+ differences were found in the medial frontal cortex and gyrus rectus including the area of the ventromedial prefrontal cortex, hippocampal areas and precuneus with posterior cingulate cortex (Supplementary Fig. S6 + S7).

Additional conjunction analyses (minimum t-statistics, conjunction null) between all six centers revealed shared activation in the bilateral insula/frontal operculum (right cluster: *k* = 415, MNI-coordinates x = 34; y = 28; z = 2, *t* = 4.74, *p_peak level_* = .039 FWE-corrected; left cluster: *k* = 500, MNI-coordinates x = −30; y = 28; z = 2, *t* = 4.74, *p_peak level_* = .321 FWE-corrected; assessed at *p*<.005 uncorrected and a cluster threshold of *k*=175) for the contrast CS+ > CS- during the first extinction phase across both measurement points (t1 + t2). No conjoint activation above cluster threshold was found for the opposite contrast CS- > CS+ during the first extinction phase.

##### Exploration of the reliability of fMRI data

According to the results in the analysis of extinction training effects, the eigenvariates from a 5 mm radius sphere around the peak voxel at the right insula (MNI-coordinates: x = 34; y = 24; z = 4), left insula (MNI-coordinates: x = −30; y = 20; z=8) and cingulate cortex (MNI-coordinates: x = 4; y = 38; z = 38) were extracted.

###### Recall of CS/US-memories

In the analysis of the test-retest-reliability of the CS+ during Ex1, we found fair ICC-values for the right insula (*ICC* = .54, *F*(99,100) = 3.31, *p* < .001), left insula (*ICC* = .57, *F*(99,100) = 3.66, *p* < .001) and middle cingulate cortex (*ICC* = .40, *F*(99,100) = 2.35, *p* < .001). In the analysis of the test-retest-reliability of the CS+ > CS- difference during Ex1, we found fair ICC-values for the right insula (*ICC* = .44, *F*(99,100) = 2.56, *p* < .001) and low ICC-values for the left insula (*ICC* = .39, *F*(99,100) = 2.30, *p* < .001) and middle cingulate cortex (*ICC* = .34, *F*(99,100) = 2.03, *p* < .001).

###### Extinction training effects

In the analysis of the test-retest-reliability of the CS+ extinction training effect (Ex1 > Ex2), we found low ICC-values for the left insula (*ICC* = .22, *F*(99,100) = 1.56, *p* = .013) and middle cingulate cortex (*ICC* = .29, *F*(99,100) = 1.81, *p* = .002), however no significant ICCs for the right insula (*ICC* = .14, *F*(99,100) = 1.33, *p* = .08). In the analysis of the test-retest-reliability of extinction training effect of the CS+ > CS- difference (Ex1 > Ex2) we found a low ICC-value for the left insula (*ICC* = .20, *F*(99,100) = 1.51, *p* = .020), however no significant ICCs for the right insula (*ICC* = .008, *F*(99,100) = 1.02, *p* = .47) or middle cingulate cortex (*ICC* = .13, *F*(99,100) = 1.3, *p* = .10).

###### Reinstatement effects

No significant ICCs for the activation difference CS+ > CS- pre reinstatement (*ICC* = .01, *F*(99,100) = 1.02, *p* = .47), post reinstatement (*ICC* = -.05, *F*(99,100) = .91, *p* = .68) or pre vs. post reinstatement (*ICC* = -.12, *F*(99,100) = .78, *p* = .89) in the cingulate cortex cluster (x = 0; y = 40; z = 10; Supplementary Fig. S3) were found.

## 4. Discussion

Fear extinction is a relevant process to understand how conditioned fear can be inhibited. It is suggested to be one key mechanism of change during exposure-based therapy in anxiety disorders. Despite the existing evidence about fear extinction and reinstatement of fear, knowledge about the actual extinction training related changes in neural correlates is sparse. Here we provide 1) evidence that extinction training is accompanied by attenuation of neural activation in regions associated with defensive reactions and fear conditioning (e.g., insula, cingulate cortex) and 2) we show that these effects are stable across time points and sites, allowing the investigation of related processes in context of clinical multi-center studies.

In line with our first hypothesis and findings from our pilot study (Hollandt et al., 2020), we found that the delayed extinction paradigm evoked strong CS+ > CS- differences in neural activation in the bilateral insular and cingulate cortex during the first extinction phase at both measurement points, supporting the hypothesis that the fear memory was successfully activated. For the CS- > CS+ contrast, we found activation in the precuneus, PCC, middle frontal gyrus and angular gyrus. The insula and dorsal anterior/middle cingulate cortex are regions consistently activated during anticipation of external threat stimuli (Fullana et al., 2018). This is an excellent premise for the investigation of extinction training related learning processes, since our paradigm targets exactly those regions expected to be involved in fear extinction learning.

On the behavioral level, at both measurement points, the re-occurrence of threat memories and conditioned responding lead to higher US-expectancy, higher arousal and lower valence regarding CS+ than CS-. Together with the findings on the behavioral level, in the SCR and startle response at day 1, this provides evidence that fear acquisition training was successful and it supports the premise that the explicit fear memory was successfully activated on a level of subjective appraisal on day 2.

Confirming our first hypothesis, effects of extinction training were reflected by a decrease of neural activation associated with the CS+ > CS- difference. The interaction between first and second extinction phase showed a distinctly reduced CS+ > CS- difference in the insular and cingulate cortex during the second extinction phase. In the CS- > CS+ difference, activation decrease was found in the precuneus and PCC. These extinction training effects indicate extinction learning of the new CS+/no-US information on a neural level.

The functions of the insula are complex; amongst others, it is part of neural circuits involved in sensory processing and emotion with strong evidence supporting its role in mediating fear and anxiety in fear conditioning (Gogolla, 2017). It seems likely that the insula detects mismatches in extinction learning: At the beginning of extinction training, due to the initial recall of the conditioned response, the mismatch between the predicted CS+/US-memory and the actual outcome is very high. Therefore, the insula activation is higher. Over the course of the training, learning occurs and predictions are less divergent from the actual outcome. Thus, less insula activation might be needed. This would support the hypothesis that the mechanism of extinction learning relies on expectancy violations in terms of prediction errors (Craske et al., 2014).

Geuter et al. (2017) provided evidence for a functional dissociation between anterior and posterior insula. While activity in the anterior insula reflects the summation of pain expectation and prediction errors resulting from unexpected pain, the posterior insula and parietal operculum employ stimulus intensity coding. We found activation reduction not only in both parts of the insula but also in the parietal operculum and supramarginal gyrus, which is part of the somatosensory association cortex. Since we used electric stimulation as US, intensity coding of a potential painful stimulus takes place. It is plausible that the re-acquisition with the US right before extinction training evoked increased activation in the insula, parietal operculum and supramarginal gyrus as an expectation of an unpleasant, possibly painful tactile stimulus and a response to the actual aversive US which then decreased until the end of the second extinction phase.

The bilateral insula is also part of the salience network together with the cingulate cortex. The salience network responds to behaviorally salient events and plays a crucial role in integrating sensory stimuli to initiate cognitive control (Chand, 2017). The dorsal anterior/middle cingulate cortex plays an important role in emotional awareness involving emotion expression and regulation (Smith et al., 2019). In our experiment, this region was consistently activated in the CS+ > CS- comparisons and showed a distinct activation reduction in CS+ > CS- differences during extinction training, possibly reflecting a decreasing need of emotional responses. In accordance with the idea of the salience network, activation reduction in insula and dorsal anterior/middle cingulate might reflect the process of the CS+ losing its predictive value for the US and thus the difference between CS+ and CS- its salience.

Fullana et al. (2018) confirmed extinction learning to be linked to a pattern frequently observed in human fMRI studies of “central autonomic network” brain activation with the anterior insular and dorsal anterior cingulate cortices as its main cortical components. Their co-activation apparently has been linked prominently to the elicitation of negative affective states, including threat-related anticipatory anxiety. Decreasing co-activation therefore might be evidence for lessening negative affect and anxiety, which is supported by our behavioral rating data. Our analysis extends these findings from immediate to delayed extinction learning, which is important with regard to modeling the processes in exposure-based CBT for patients with anxiety disorders.

On the behavioral level, effects of extinction training were reflected by a decreased CS+ > CS- difference in US-expectancy and arousal reduction toward the CS+ over the course of the experiment. These changes in ratings suggest extinction learning on an explicit level of cognitive and emotional appraisal. Additionally, extinction training effects in valence ratings were correlated with decreasing CS+ > CS- difference in insula and cingulate cortex. This underlies the biological foundation of psychological phenomena in learning processes.

A general activation reduction from the second extinction to the reinstatement test phase regarding the CS+ > CS- differences was found in the anterior insular and anterior cingulate cortex, suggesting an overall ongoing extinction learning where the CS+ loses its predictive value and trait as a threat for the US. The reduction of deactivation in the hippocampus and pre-/postcentral gyrus on the CS+ regarding the CS- > CS+ contrast suggests an involvement of memory as well as senso-motoric processes in a later phase of extinction training.

However, confirming our second hypothesis, a reinstatement of conditioned fear after exposure to the US post extinction training was reflected by temporary changes in neural activation and subjective appraisal. On the neural level, we found an interaction of CS-type X pre/post reinstatement in the ACC. Before the reinstatement, activation was higher during CS-presentation than during CS+, whereas after the reinstatement, this differentiation showed the opposite pattern. This activation change was furthermore positively correlated with changes in CS+ > CS- difference US-expectancy ratings, pointing again to the biological foundation of psychological phenomena. The presentation of the US alone between the end of the second extinction phase and start of the post-reinstatement phase led to a higher US-expectancy and arousal toward the CS+, which diminished fast in the course of the last extinction training phase.

In neither variable of the analysis were the CS+ > CS- differences completely vanished. This goes along with an observation from the clinical context: extinction of well-consolidated pathological fear is challenging and needs multiple exposure-based CBT sessions. Although our participants did not suffer from pathological fear, the combination of acquisition training and 24h of consolidation generated a very robust excitatory fear response. The meta-analysis by Fullana et al. (2018) confirmed that fear responses to CS+ vs. CS− are rarely completely diminished during extinction learning. On the one hand, this might be due to an insufficient number of stimulus presentations during the experiment. On the other hand, memory processes play a crucial role during extinction learning and hence traces of extinction memory need consolidation time. Besides that, it also suggests that despite successful extinction training, a residual fear caused by the original CS+/US link remains.

Furthermore, in line with our third hypothesis, we could substantially extend initial findings from Hollandt et al. (2020) and confirm the stability of neural and behavioral data associated with extinction learning processes across two measurement points and six investigation sites. We were able to find comparable effects at all sites, respectively in basic comparisons regarding neural and behavioral data as well as overall comparable effects at t1 and t2, regarding extinction training. Neither on the neural activation in fear related brain areas nor in the behavioral data we found systematic time-depended or site-dependent influences. The only overlap between site effects and CS- > CS+ extinction effects from Ex2 to RIT was found in a cluster of the occipital cortex.

Regarding the feasibility of the multicenter approach of our delayed extinction paradigm, we were able to demonstrate that it works exactly as intended repeatedly at different sites with different MRI-scanners and personnel, in site specific small samples (*n* ∼ 16). Thus, this paradigm is useful and valuable for clinical comparisons of patients before vs. after exposure-based CBT and patients compared to healthy subjects. Future studies using this delayed extinction paradigm have the potential of shedding new light on the key mechanisms of exposure-based therapy and therefore foster patient care.

In light of the recent “replication crisis” in psychology and neuroscience (Ioannidis, 2005; Open Science, 2015), task-based fMRI is harshly criticized for having poor reliability (Bennett & Miller, 2010). One measure for reliability is calculating Intraclass Correlation Coefficients (ICCs) which are particularly difficult to deal with in fMRI-settings since ICCs can be analyzed for specific regions of interest vs. whole-brain as well as different contrasts of interest, and thus produce very different outcomes (Brandt et al., 2013). Noteworthily, despite a high consistency in the activation pattern of CS+ vs. CS- effects over two measurement points and six sites on group-level, we only found low to fair ICCs for activity in clusters of the insula and cingulate cortex for selected comparisons regarding the recall of CS/US-memories and low ICCs regarding extinction training effects. This is partially lower than the ICCs we found for the behavioral data: fair ICCs for the recall of CS/US-memories and low to good ICCs for extinction training effects. Overall, low ICCs challenge and set limitations to the use of fMRI data in predicting and treating anxiety disorders at the individual level. However, low ICCs seem to be a general phenomenon in task-fMRI studies as findings from a current meta-analysis (Elliott et al., 2020) of 90 fMRI experiments (n=1,008) across 11 common fMRI tasks revealed a mean ICC of .397. The authors discuss their findings by explaining that task-fMRI (as well as most behavioral) paradigms are intentionally designed to reveal how the average human brain responds to provocation, while minimizing between-subject variance. They did not find evidence that MRI measures, or even the BOLD signal itself, are inherently unreliable. But the majority of task-fMRI measures are based on contrasts between conditions and explicitly not between individuals, which is also true for our fear conditioning experiment.

On the one hand, it might be that a first extinction training is not comparable with a second timepoint (i.e., high within-subject variability) due to the dynamic nature of learning paradigms, which shapes organisms to individually adapted to their environment. Considered this, ICCs up to .67 show that there is at least some accordance across time points. On the other hand, another possible explanation for low ICCs in experimental paradigms in general could be relatively low between-subject variability due to high homogeneity in the sample (plausible in our case, since we only investigated healthy subjects) in relation to the within-subjects variability. As a result, robust (different groups at different scanners) and replicable (the same group at multiple measurement points) paradigms (comparable result patterns) on group-level exist, although having low inter-individual reliability. These paradigms can therefore be useful for the investigation of between-group differences in the first place (Hedge et al. 2018). Analyses of the individual trajectories on the basis of our paradigm, however, might profit from different approaches than the traditional group- and event-contrasting fMRI analyses – e.g., machine learning (Hahn et al., 2015) – as suggested by Elliott et al. (2020). Considering that we explored ICCs on the basis of activation observed in difference contrasts on group level, this might be a confounding issue here.

Although we carefully conducted our study and analyses, several limitations must be considered. To test our hypothesis about extinction training and reinstatement, we used the power of the large sample across both measurement points, while for the exploration of site and time effects we only tested the comparisons CS+ > CS- and CS- > CS+ at the beginning of the experiment in which effect sizes were expected to be largest. Based on our results, we can only assume that extinction training related decrease in CS+ vs. CS- activation differences as we found in the combined sample would also be found at each site given a larger sample.

As anticipated, the consolidated conditioned reactions in form of higher US-expectancy, higher arousal, lower valence and higher neural activation in the insula and cingulate cortex towards the CS+ compared to the CS- was quite strong and extinction training effects rather small. The rating data showed that even at the very end of the experiment, there were still significant differences between CS+ and CS- ratings. Presumably, longer extinction training would be necessary to completely diminish differences in the reaction towards CS+ and CS-. However, since we are highly interested in investigating patients with anxiety disorders with this paradigm, we had to make the practical decision to cut back on MRI scanning time. The MRI environment with its very limited space, no possibility to move, and loud noises can be a stressful experience even for healthy subjects but even more so for patients who suffer from anxiety. Therefore, we compromised the length of the experiment so that it was manageable for patients.

Regarding the ICC reliability analyses we had to face the challenge of a multicenter study that was not initially designed to test the reliability of our paradigm. Ideally, all participants are tested multiple times on the same set of different MRI-scanners so that test-retest reliability as well as inter-scanner reliability can be disentangled (Friedman et al., 2008). Since we were only interested in the overall quality and reproducibility of our paradigm and neither looking for definitive biomarkers nor individual differences research, we focused on exploring the regions our extinction training targets in line with the current literature (Fullana et al. 2018). Also, we only tested each participant twice in the same scanner. Therefore, we had to remove the influence of site by regressing the eigenvariates to site and only explore the test-retest reliability of the residuals. For a proper investigation of our paradigm’s reliability, a specifically planned study would be needed.

In conclusion, we could validate our delayed extinction fMRI-paradigm to be suitable for the investigation of fear extinction. As a necessary further development in extinction research, we specifically focused on process information of neural correlates associated with fear extinction training and reinstatement to shed new light on the possible neural mechanisms underlying exposure-based CBT and could show convergent findings in behavioral and neural data.

Furthermore, we provide evidence that our paradigm is robust and appropriate for multicentric settings. Thus, it provides a solid basis for future clinical investigations (pre/post exposure therapy) on patients with anxiety disorders as it is already in preparation in the PROTECT-AD consortium.

## Disclosures and acknowledgments

### Staff members by site

Members of the consortium are: Dresden: Hans-Ulrich Wittchen, Ingmar Heinig, Andre Pittig, Lucie Scholl, Katrin Hummel, Dorte Westphal. Bochum: Jürgen Margraf, Silvia Schneider, Verena Pflug, Christina Totzeck, Dirk Adolph, Cornelia Mohr, Jan Cwik. Greifswald: Alfons Hamm, Jan Richter, Maike Hollandt, Anne Pietzner, Jörg Neubert. Marburg: Benjamin Straube, Tilo Kircher, Carsten Konrad, Yunbo Yang, Isabelle Ridderbusch, Hanna Christiansen, Winfried Rief, Katrin Wambach, Anne Maenz, Sophia Tennie. Würzburg: Jürgen Deckert, Katharina Domschke, Paul Pauli, Marcel Romanos, Ulrike Lueken, Kathrin Zierhut Kristina Dickhöver, Markus Winkler, Maria Stefanescu, Christiane Ziegler. Berlin: Andreas Ströhle, Thomas Fydrich, Isabel Alt, Sophie Bischoff, Jennifer Mumm. Münster: Volker Arolt, Katja Koelkebeck, Udo Dannlowski, Nathalia Weber, Sebastian Schauenberg, Sophia Wriedt. The project was carried out in conjunction with Xina Grählert and Marko Käppler of the Coordinating Centre for Clinical Trials (KKS) and Michael Höfler and Jens Strehle of the Centre for Clinical Epidemiology and Longitudinal Studies (CELOS) of Technische Universität Dresden.

### Data Access and Responsibility

All principle investigators take responsibility for the integrity of the respective study data and their components. All authors and co-authors had full access to all study data. Data analysis and manuscript preparation were completed by the authors and co-authors of this article, who take responsibility for its accuracy and content.

### Ethical standards

The study program is performed according to the Declaration of Helsinki and was approved by the Ethics Committee of Technische Universität Dresden (EK 234062014, November 14, 2014). The clinical trial in adults has been registered with NIMH Protocol Registration System (01EE1402A) and with the German Register of Clinical Studies (DRKS00008743).

## Data Availability

Due to privacy restrictions of the collected data, we are not able to make the raw data openly available. However, group level data will be made available at NeuroVault upon acceptance of the manuscript.

