## Supplemental Material for "Neural adaptation of cingulate and insular activity during delayed fear extinction: A replicable pattern across assessment sites and repeated measurements"

### Supplement

#### 1. Methods

##### *1.1. Quality control of MRI data-sets*

The PROTECT-AD add-on project “neural response and fear circuitry related to extinction learning and outcome”, funded by the German Federal Ministry of Education and Research (BMBF) was carefully planned and monitored. MRI sequences and the paradigm were tested in a pilot study (Hollandt et al., 2020). An identical setup for electrical dermal stimulation in the lab (day 1) and the MRI environment (day 2) had been implemented at each site. The sequences and the scanner comparability were further evaluated before the start and across the entire data-acquisition phase by applying the same measurement sequences to MRI phantoms. A detailed study protocol was developed to obtain a homogeneous data sample. The data-acquisition was further monitored and supervised by monthly telephone conferences and site visits.

In total, n = 119 healthy subjects as controls for patients with anxiety disorders were enrolled in the PROTECT-AD add-on project. Besides the extinction paradigm a resting state paradigm as well as an emotional face recognition paradigm that are not part of the analyses of this manuscript, were applied. Initially, data was collected at seven sites in Germany: n = 17 at site 1, n = 10 at site 2, n = 17 at site 3, n = 15 at site 4, n = 15 at site 5, n = 30 at site 6, n = 15 at site 7.

The first inclusion criterion for the current investigation was data completeness of the extinction paradigm for both measurement points (t1 and t2). N = 9 participants were excluded because at least one (t1 or t2) extinction data-set was missing. These n = 110 data-sets were composed of n = 15 at site 1, n = 7 at site 2, n = 16 at site 3, n = 14 at site 4, n = 15 at site 5, n = 28 at site 6, n = 15 at site 7. From the remaining n = 110 complete (t1 + t2) data-sets, another n = 10 were excluded due to the following criteria: artifacts in the visual inspection, percent signal change (PSC; Stöcker et al., 2005), or head-movement > 1,5 voxel sizes (6 mm).

Visual inspection of the raw- and pre-processed data was performed with special regard to extinctions, susceptibility artefacts, other more global aspects of head shape or visible artifacts (lines in scans) as well as artefacts in and extinctions of regions around the amygdala. 6 data-sets had to be excluded (n = 5 from site 2, n = 1 from site 6)

The PSC was calculated for all data sets to exclude data with deviants in the signal fluctuation. By considering PSC with a cut-off threshold of 2.5 standard deviations of the mean, 2 data-sets had to be excluded (n = 2 from site 2).

Head movement was set at 1.5 voxel sizes to exclude data sets (x, y or x axis > 6 mm) for at least one measurement point (t1 or t2). Two data-sets had to be excluded (n = 1 from site 6, n = 1 from site 7).

Taken together, this eventually resulted in a quality-controlled sample of n = 100 composed of: n = 15 at site 1, n = 0 at site 2, n = 16 at site 3, n = 14 at site 4, n = 15 at site 5, n = 26 at site 6, n = 14 at site 7. Figure S1 is a visualization of the quality control process.

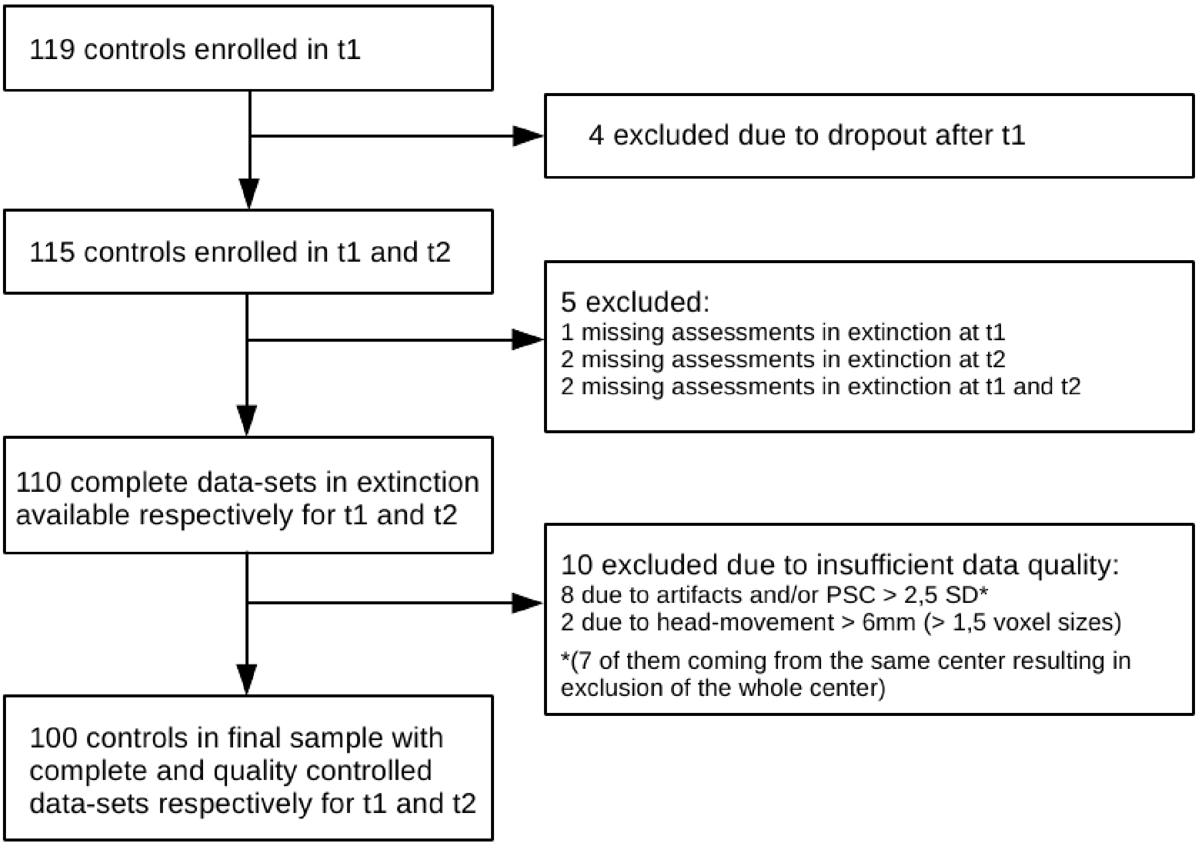


**Figure S1.** Visualization of the quality control process.

*1.2. Details of experimental procedures (day 1+2)*

As described in Hollandt et al. (2020), two background-colored pictures of male faces with neutral expressions (from the Psychological Image Collection at Stirling; http://pics.stir.ac.uk, following Duits et al., 2017) served as CSs (counterbalanced between subjects). CSs were presented for 6000 ms, followed by an inter-trial interval (white fixation cross presented on a black screen) of 6 to 10 s. An unpleasant electrocutaneous stimulus was used as US with a stimulus duration of 500 ms, composed of 100 single pulses of 5 ms. It was presented after 5.5 s after stimulus onset, so that it co-terminated with the CS+. For the application, a constant current stimulator (DS7A, Digitimer, Medical Products, Wiesbaden) with MRI compatible reusable cup electrodes (10 mm silver, Medical Products, Wiesbaden) and specially produced electrode gel were used. The electric stimuli were triggered by the presentation software and delivered to the forearm of the non-dominant hand. For the extinction training, current intensity and electrode position were adopted from the adjustments of the first day.

Using the Presentation 17.2 software (Neurobehavioral Systems, Albany, CA, http://www.neurobs.com/), all stimuli inside the MR scanner were presented on an MRI-compatible LCD screen, visible through a mirror attached to the MR head coil.

Due to technical limitations inside the scanner an 11-point scale for the US expectancy rating had to be used allowing ratings in steps of 10%. Furthermore, the subjects had only 5 s for the rating to shorten the length of the fMRI paradigm.

To avoid sequence effects, two orders were used (randomized between subjects) differing in the sequence of stimulus presentations each beginning with one of the two CS. The CS presentation followed a pseudo-randomized order with no more than two consecutive presentations of the same CS.

*1.3 Procedure details on psychophysiological assessment during fear conditioning (day 1)*

At day 1, a 50 ms burst of white noise with an intensity of 95 dB[A] (rise/fall < 1ms) served as a startle-eliciting probe stimulus and was presented binaurally over Sennheiser AKG K66 headphones either 4.5 or 5 sec after CS onset and during the ITI (2, 3, 4, 5, or 6sec after CS offset). Startle probes were presented during all CS trials during pre-conditioning phase and during 8 of 10 CS trials each during fear acquisition trainings, and twice as often during the respective ITI. Before pre-conditioning, 6 startle probes were presented alone to ensure a stable baseline of blink magnitudes.

The eyeblink component of the startle reflex was measured by recording electromyographic (EMG) activity over the orbicularis oculi muscle beneath the left eye, using two Ag/AgCl surface electrodes (4mm diameter, F-E9-60, Warwick, RI), filled with electrolyte paste (GE Medical Systems Milwaukee, WI). The EMG raw signal was amplified and filtered through a 30 Hz to 1000 Hz bandpass filter using a Coulbourn S75-01 bioamplifier (Allentown, PA) and a 400 Hz Kemo-VFB-8-03 low pass filter (Kemo, Dartford, UK). The digital sampling with a rate of 1000 Hz started 100ms before and lasted until 100ms after the onset of the acoustic startle probe. The EMG signal was filtered offline using a 60 Hz highpass filter and was rectified and integrated (time constant: 10ms) by a digital filter.

To record skin conductance from the hypothenar muscle of the palmar surface of the participants' non-dominant hand (Boucsein et al., 2012), a Coulbourn S71-22 skin conductance coupler was used, providing a constant 0.5 V across two Ag/AgCl surface electrodes (8 mm diameter, E224A, Warwick, RI; filled with isotonic 0.5 M sodium chloride electrode gel). The sampling rate was 10 Hz.

*Data reduction and response definition*

*Skin conductance response:* The skin conductance responses were scored as the first response in conductance within a 0.90 - 4.00 s time window, following stimulus onset for CS and US, respectively, using an in-house program (Globisch et al., 1993). Trials in which no SCR could be detected were scored as zero responses. Missing values were replaced individually for each subject by the overall SCR of this subject over all trials of the respective stimulus during the experiment. Logarithms for each value were then computed to normalize the distribution (Venables & Christie, 1980). To reduce interindividual variability of the SCR that was not related to the conditioning and extinction tasks of the experiment, the log values were range-corrected (division of individual score by the participants' maximum response within all CS and US trials (Lykken & Venables, 1971)).

*Startle blink magnitudes:* Data were semi-automatically scored offline by using an in-house algorithm (Globisch et al., 1993), that identified latency of blink onsets and peak amplitudes. The time window for startle response was defined between 20-120ms after startle probe onset and the magnitude had to peak within 150 ms after onset. No detectable eyeblinks were scored as zero responses. Trials with excessive baseline activity, recording artifacts (e.g., electrode malfunctions), and spontaneous eyeblinks outside the latency window were treated as missings and therefore rejected. All participants met the 80% criterion for valid responses and could be included in statistical analysis. The missing values were replaced individually for each subject by the overall mean blink response magnitude of this subject over all trials of the experiment. Each response of each participant was then standardized and converted to T-Scores [50+($z \times 10$)] to control for possible confounding effects of high inter-individual differences in baseline amplitude.

Physiological and rating data were reduced by averaging two trials (or in case of blink magnitudes two probes) to one block.

Due to missing data caused by technical failures, quality-controlled data for ratings, startle blink magnitude and skin conductance responses were available only in n = 90, n = 75, and n = 66 subjects, respectively.

All conditioning data were analyzed using repeated-measures ANOVAs with Stimulus (CS+ vs. CS−) and Block as within-subject factors. All tests were conducted two-sided and uncorrected for multiple comparisons. A Greenhouse–Geisser procedure was used in case of a violation of the sphericity assumption in ANOVAs. $\eta^2$ values are provided as a measure of effect size. Results are described in the Supplementary Results 2.1.

#### 2. Results

*2.1 Conditioning on day 1, see Supplementary Figure S2*

*US-expectancy:* At t1, prior to any conditioning, there was no difference between CS+ and CS- in CS/US-expectancy (t1: $F(1,89) = 1.24$, $p = .27$, $\eta_{p^2} = .014$; t2: $F(1,89) = 7.91$, $p = .006$, $\eta_{p^2} = .08$).

At both measurement points, over the course of the experiment, the CS+/US-expectancy increased (t1: $F(1,4) = 9.72$, $p < .001$, $\eta_{p^2} = .10$; t2: $F(1,4) = 18.13$, $p < .001$, $\eta_{p^2} = .17$), whereas the CS-/US-expectancy decreased (t1: $F(1,4) = 4.24$, $p = .014$, $\eta_{p^2} = .05$; t2: $F(1,4) = 9.91$, $p < .001$, $\eta_{p^2} = .10$).

After acquisition, there was a distinct difference between CS+ and CS- in US-expectancy rating, indicating that conditioning was successful (t1: $F(1,89) = 1221.34$, $p < .001$, $\eta_{p^2} = .93$; t2: $F(1,89) = 1413.63$, $p < .001$, $\eta_{p^2} = .94$).

*Arousal:* At both measurement points, prior to any conditioning, there was no difference between CS+ and CS- in arousal (t1: $F(1,89) = .15$, $p = .70$, $\eta_{p^2} = .002$; t2: $F(1,89) = 3.88$, $p = .05$, $\eta_{p^2} = .04$).

After acquisition, there was a distinct difference between CS+ and CS- arousal rating, indicating that conditioning was successful (t1: $F(1,89) = 108.64$, $p < .001$, $\eta_{p^2} = .55$; t2: $F(1,89) = 126.04$, $p < .001$, $\eta_{p^2} = .59$).

*Valence:* At both measurement points, prior to any conditioning, there was no difference between CS+ and CS- in valence (t1: $F(1,89) = 1.25$, $p = .27$, $\eta_{p^2} = .014$; t2: $F(1,89) = .18$, $p = .68$, $\eta_{p^2} = .002$).

After acquisition, there was a distinct difference between CS+ and CS- valence rating, indicating that conditioning was successful (t1: $F(1,89) = 56.49$, $p < .001$, $\eta_{p^2} = .39$; t2: $F(1,89) = 62.13$, $p < .001$, $\eta_{p^2} = .41$).

*SCR:* At both measurement points, prior to any conditioning, there was no difference between CS+ and CS- in SCR (t1: $F(1,65) = .38$, $p = .54$, $\eta_{p^2} = .006$; t2: $F(1,65) = .28$, $p = .60$, $\eta_{p^2} = .004$).

At the start of conditioning, there was a significant difference between CS+ and CS- in SCR (t1: $F(1,65) = 53.88$, $p < .001$, $\eta_{p^2} = .45$; t2: $F(1,65) = 43.41$, $p < .001$, $\eta_{p^2} = .40$), which (despite some habituation) was still present at the end of conditioning (t1: $F(1,65) = 20.91$, $p < .001$, $\eta_{p^2} = .24$; t2: $F(1,65) = 6.41$, $p = .014$, $\eta_{p^2} = .09$).

*Startle:* At both measurement points, prior to any conditioning, there was no difference between CS+ and CS- in startle blink magnitude (t1: $F(1,74) = .36$, $p = .55$, $\eta_{p^2} = .005$; t2: $F(1,74) = .09$, $p = .76$, $\eta_{p^2} = .001$).

At the start of conditioning, there was a distinct difference between CS+ and CS- in startle blink magnitude (t1: $F(1,74) = 23.72$, $p < .001$, $\eta_{p^2} = .24$; t2: $F(1,74) = 38.63$, $p < .001$, $\eta_{p^2} = .34$), which (despite habituation processes) was still present at the end of conditioning (t1: $F(1,74) = 30.37$, $p < .001$, $\eta_{p^2} = .29$; t2: $F(1,74) = 27.75$, $p < .001$, $\eta_{p^2} = .27$).

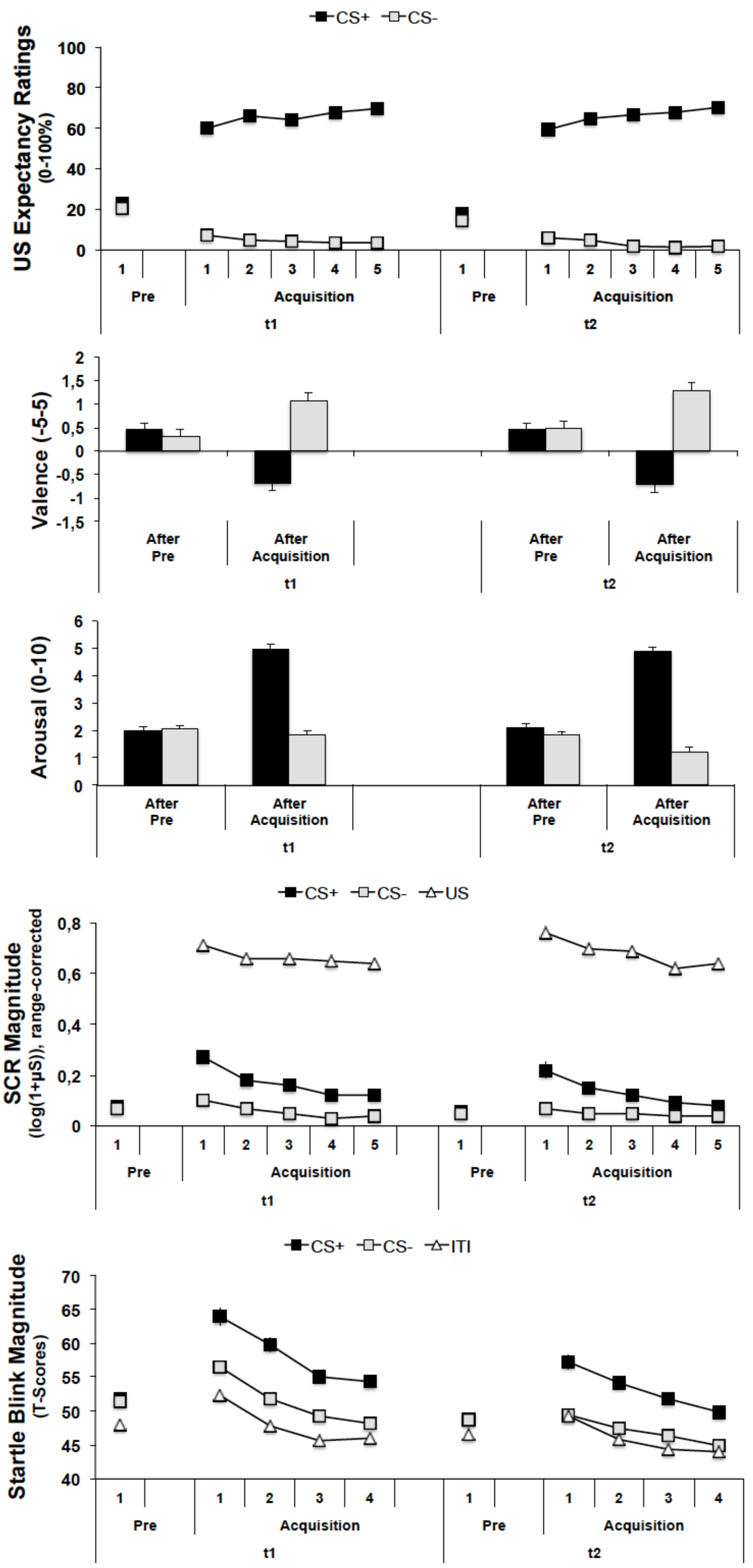


**Figure S2.** Mean Scores and standard errors for US expectancy ratings, ratings of valence and arousal, SCRs and startle blink magnitudes, respectively, during phases of pre-conditioning and fear acquisition (day 1) at t1 and t2 as a function of stimulus type (CS+ and CS-, as well as US and ITI in case of SCR and startle, respectively) with two trials per block for continuously assessed measures.

*2.2 Recall of CS/US-associations in behavioral data:*

*US-expectancy:* At both measurement points, the recall of CS/US-associations learned the day before during acquisition training prior to any extinction training and prior to the re-acquisition of CS+ and US showed a higher US-expectancy during CS+ than during CS- (t1: $F(1,98) = 80.31$, $p < .001$, $\eta_{p^2} = .45$; t2: $F(1,98) = 86.96$, $p < .001$, $\eta_{p^2} = .47$).

*Arousal:* At both measurement points, prior to any extinction training, there was higher arousal regarding the CS+ than the CS- (t1: $F(1,98) = 67.93$, $p < .001$, $\eta_{p^2} = .41$; t2: $F(1,98) = 88.96$, $p < .001$, $\eta_{p^2} = .48$).

*Valence:* At both measurement points, prior to any extinction learning, there was lower valence regarding the CS+ than the CS- (t1: $F(1,98) = 16.70$, $p < .001$, $\eta_{p^2} = .15$; t2: $F(1,98) = 51.85$, $p < .001$, $\eta_{p^2} = .35$).

*2.3 Effect of re-acquisition in behavioral data*:

*US-expectancy:* After re-acquisition, the CS+/US-expectancy was increased (t1: $F(1,98) = 30.45$, $p < .001$, $\eta_{p^2} = .24$; t2: $F(1,98) = 26.61$, $p < .001$, $\eta_{p^2} = .21$) whereas the CS-/US-expectancy was decreased (t1: $F(1,98) = 3.46$, $p = .06$, $\eta_{p^2} = .03$; t2: $F(1,98) = 17.63$, $p < .001$, $\eta_{p^2} = .15$).

**Table S1.** Activation difference CS+ vs. CS- during first extinction phase at both measurement points (t1, t2). All contrasts were assessed at *p*<.005 uncorrected with a cluster threshold of *k*=175.

| Region | hemi-sphere | no. voxels | MNI coordinates peak x | y | z | t | P-value FWE-corrected peak | cluster |
|---|---|---|---|---|---|---|---|---|
| **t1: CS+ > CS-** | | | | | | | | |
| Anterior insula + frontal operculum | R | 8348 | 34 | 24 | 4 | 10.84 | <.001 | <.001 |
| Parietal operculum + supramarginal gyrus + superior temporal gyrus | L | 1665 | -56 | -26 | 24 | 9.31 | <.001 | <.001 |
| Supramarginal gyrus + angular gyrus + parietal operculum | R | 2866 | 60 | -42 | 30 | 7.47 | <.001 | <.001 |
| Middle cingulate cortex + supplementary motor cortex + superior frontal gyrus medial segment | | 6873 | 0 | 18 | 36 | 7.41 | <.001 | <.001 |
| Precentral gyrus + middle frontal gyrus | R | 1833 | 42 | 0 | 48 | 6.34 | <.001 | <.001 |
| Cerebellum | R | 738 | -32 | -52 | -28 | 5.52 | .001 | .004 |
| Precuneus | L | 384 | 12 | -64 | 40 | 5.38 | .002 | .087 |
| Precentral gyrus | L | 189 | -40 | -4 | 48 | 5.04 | .009 | .530 |
| Middle + superior frontal gyrus | R | 264 | 52 | -30 | -6 | 4.74 | .033 | .270 |
| Middle frontal gyrus | L | 619 | -36 | 40 | 36 | 4.36 | .143 | .011 |
| Cerebellum | R | 226 | 32 | -52 | -28 | 4.32 | .162 | .384 |
| Cerebellum | | 274 | 0 | -50 | -20 | 4.10 | .331 | .246 |
| **t1: CS- > CS+** | | | | | | | | |
| Precuneus + posterior cingulate cortex + fusiform gyrus + hippocampus | R | 3293 | 2 | -54 | 26 | 7.31 | <.001 | <.001 |
| Angular gyrus + superior parietal lobule | L | 3684 | -48 | -68 | 30 | 6.96 | <.001 | <.001 |
| Middle + superior temporal gyrus | L | 503 | -60 | -12 | -16 | 6.40 | <.001 | .030 |
| Medial frontal cortex + superior frontal gyrus medial segment + superior frontal gyrus | R | 2082 | 2 | 40 | -18 | 6.37 | <.001 | <.001 |
| Angular gyrus + occipital gyrus | R | 4733 | 52 | -66 | 28 | 5.82 | <.001 | <.001 |
| Precentral gyrus | L | 3308 | -10 | -28 | 74 | 5.15 | .005 | <.001 |
| Postcentral gyrus | R | 1134 | 62 | -4 | 20 | 4.83 | .020 | <.001 |
| Superior + middle frontal gyrus | R | 231 | 20 | 36 | 46 | 4.03 | .389 | .367 |
| **t2: CS+ > CS-** | | | | | | | | |
| Anterior insula + frontal + central operculum | R | 4336 | 36 | 24 | 2 | 11.47 | <.001 | <.001 |
| Anterior insula + frontal + central operculum | L | 2536 | -32 | 20 | 6 | 9.74 | <.001 | <.001 |
| Middle cingulate gyrus + superior frontal gyrus medial segment + supplementary motor cortex | R | 6144 | 4 | 36 | 40 | 8.91 | <.001 | <.001 |
| Supramarginal gyrus + angular gyrus + parietal operculum | R | 2913 | 60 | -42 | 34 | 7.70 | <.001 | <.001 |
| Supramarginal gyrus + postcentral gyrus + parietal operculum | L | 1204 | -62 | -24 | 26 | 7.01 | <.001 | <.001 |
| Precentral gyrus + middle fontral gyrus | R | 872 | 44 | 2 | 44 | 5.82 | <.001 | <.001 |
| Precentral gyrus + middle fontral gyrus | L | 234 | -38 | -4 | 46 | 5.16 | .001 | .001 |
| Cerebellum | L | 445 | -16 | -76 | -28 | 4.68 | .003 | <.001 |
| Middle frontal gyrus | R | 686 | 38 | 48 | 16 | 4.53 | .077 | .006 |
| Middle frontal gyrus | L | 292 | -34 | 52 | 24 | 4.18 | .259 | .207 |
| t2: CS- > CS+ | | | | | | | | |
| Angular gyrus + precuneurs + posterior cingulate gyrus + precentral gyrus medial segment | L | 29998 | -40 | -68 | 32 | 8.82 | <.001 | <.001 |
| Middle frontal gyrus + precentral gyrus + opercular part of the inferior frontal gyrus | L | 793 | -46 | 26 | 22 | 5.28 | .003 | .003 |

**Abbreviations:** CS+: conditioned stimulus that is followed by the unconditioned stimulus (US) with a reinforcement rate of 60% (only unpaired CS+ were included; CS-: conditioned stimulus that is never followed by an US; L: left; R: right; no. voxel: number of voxels per cluster; x, y, z: MNI coordinates.

**Table S2. A)** 3-way ANOVAs with CS-type (CS+, CS-), Block (Pre Re-Acquisition, Post Re-Acquisition, Post Ex1, Post Ex2, Post Reinstatement, Post Ex3) and Time (t1, t2) as within-subject factors. **B)** Rating comparisons between first (t1) and second (t2) point measurement regarding CS+ > CS- differences. Data were analyzed using repeated-measures ANOVAs with Measurement (t1, t2) as within-subjects.

| **A) 3-way ANOVA** | | | | |
|---|---|---|---|---|
| **US-expectancy** | **dfs** | **F** | **P** | $\eta_p^2$ |
| Time | 1, 98,00 | 8.52 | .004 | .080 |
| Block | 1, 98,00 | 74.46 | < .001 | .432 |
| CS | 1, 98,00 | 301.33 | < .001 | .760 |
| Time x Block | 1, 98,00 | 2.96 | .031 | .029 |
| Time x CS | 1, 98,00 | .30 | .58 | .003 |
| Block x CS | 1, 98,00 | 20.19 | < .001 | .171 |
| Time x Block x CS | 1, 98,00 | 1.15 | .33 | .012 |
| **Arousal** | | | | |
| Time | 1, 98,00 | 5.18 | .03 | .050 |
| Block | 1, 98,00 | 48.28 | < .001 | .330 |
| CS | 1, 98,00 | 135.50 | < .001 | .580 |
| Time x Block | 1, 98,00 | 3.56 | .01 | .035 |
| Time x CS | 1, 98,00 | 3.36 | .07 | .033 |
| Block x CS | 1, 98,00 | 6.79 | < .001 | .065 |
| Time x Block x CS | 1, 98,00 | .35 | .85 | .004 |
| **Valence** | | | | |
| Time | 1, 98,00 | .61 | .44 | .006 |
| Block | 1, 98,00 | 6.38 | < .001 | .061 |
| CS | 1, 98,00 | 62.36 | < .001 | .389 |
| Time x Block | 1, 98,00 | 1.15 | .33 | .012 |
| Time x CS | 1, 98,00 | 4.55 | .04 | .044 |
| Block x CS | 1, 98,00 | 3.65 | .01 | .036 |
| Time x Block x CS | 1, 98,00 | .69 | .60 | .007 |
| **B) Comparisons between t1 and t2 regarding CS+ > CS- differences** | | | | |
| **US-expectancy t1 vs. t2** | **dfs** | **F** | **P** | $\eta_p^2$ |
| Pre Re-Acquisition CS+ > CS- | 1, 98,00 | .12 | .73 | .001 |
| Post Re-Acquisition CS+ > CS- | 1, 98,00 | 1.46 | .23 | .015 |
| Post Ex1 CS+ > CS- | 1, 98,00 | .41 | .52 | .004 |
| Post Ex2 CS+ > CS- | 1, 98,00 | 2.11 | .15 | .021 |
| Post Reinstatement CS+ > CS- | 1, 98,00 | .45 | .50 | .005 |
| Post Ex3 CS+ > CS- | 1, 98,00 | .70 | .41 | .007 |
| **Arousal t1 vs. t2** | | | | |
| Pre Ex1 CS+ > CS- | 1, 98,00 | 1.23 | .27 | .013 |
| Post Ex1 CS+ > CS- | 1, 98,00 | 1.84 | .18 | .018 |
| Post Ex2 CS+ > CS- | 1, 98,00 | 3.89 | .051 | .038 |
| Post Reinstatement CS+ > CS- | 1, 98,00 | .27 | .61 | .003 |
| Post Ex3 CS+ > CS- | 1, 98,00 | 2.59 | .11 | .026 |
| **Valence t1 vs. t2** | | | | |
| Pre Ex1 CS+ > CS- | 1, 98,00 | 3.77 | .06 | .037 |
| Post Ex1 CS+ > CS- | 1, 98,00 | 3.53 | .06 | .035 |
| Post Ex2 CS+ > CS- | 1, 98,00 | 2.01 | .16 | .020 |
| Post Reinstatement CS+ > CS- | 1, 98,00 | 3.57 | .06 | .035 |
| Post Ex3 CS+ > CS- | 1, 98,00 | .64 | .43 | .006 |

**Table S3.** Basic contrasts in ratings at each site separately.

| **US-expectancy** | | | | |
|---|---|---|---|---|
| *t1+t2 CS+ > CS- post re-acquisition* | **df** | **F** | **P** | $\eta_p^2$ |
| Site 1 | 1, 14 | 46.98 | <.001 | .770 |
| Site 2 | 1, 15 | 37.20 | <.001 | .713 |
| Site 3 | 1, 13 | 168.66 | <.001 | .928 |
| Site 4 | 1, 14 | 133.41 | <.001 | .905 |
| Site 5 | 1, 25 | 68.38 | <.001 | .732 |
| Site 6 | 1, 12 | 28,74 | <.001 | .705 |
| *t1+ t2 CS+ extinction post re-acquisition vs. post Ex1 vs. post Ex2* | | | | |
| Site 1 | 2, 14 | 13.79 | <.001 | .496 |
| Site 2 | 2, 15 | .84 | .44 | .053 |
| Site 3 | 2, 13 | 24.02 | <.001 | .649 |
| Site 4 | 2, 14 | 9.65 | .001 | .408 |
| Site 5 | 2, 25 | 36.20 | <.001 | .695 |
| Site 6 | 2, 12 | 5.12 | .014 | .299 |
| **Arousal** | | | | |
| *t1+t2 CS+ > CS- pre Ex1* | | | | |
| Site 1 | 1, 14 | 14.08 | .002 | .501 |
| Site 2 | 1, 15 | 14.55 | .002 | .492 |
| Site 3 | 1, 13 | 37.43 | <.001 | .742 |
| Site 4 | 1, 14 | 21.08 | <.001 | .601 |
| Site 5 | 1, 25 | 16.22 | <.001 | .393 |
| Site 6 | 1, 12 | 20.17 | .001 | .627 |
| *t1+ t2 CS+ extinction pre Ex1 vs. post Ex1 vs. post Ex2* | | | | |
| Site 1 | 2, 14 | 2.26 | .12 | .139 |
| Site 2 | 2, 15 | 2.62 | .089 | .149 |
| Site 3 | 2, 13 | 8.77 | .001 | .403 |
| Site 4 | 2, 14 | .45 | .65 | -031 |
| Site 5 | 2, 25 | 14.81 | <.001 | .372 |
| Site 6 | 2, 12 | 7.05 | .004 | .370 |
| **Valence** | | | | |
| *t1+t2 CS+ > CS- pre Ex1* | | | | |
| Site 1 | 1, 14 | 8.41 | .012 | .375 |
| Site 2 | 1, 15 | 12.02 | .003 | .445 |
| Site 3 | 1, 13 | 16.81 | .001 | .564 |
| Site 4 | 1, 14 | 16.44 | .001 | .540 |
| Site 5 | 1, 25 | 9.35 | .005 | .272 |
| Site 6 | 1, 12 | 7.67 | .017 | .390 |
| *t1+ t2 CS+ extinction pre Ex1 vs. post Ex1 vs. post Ex2* | | | | |
| Site 1 | 2, 14 | .83 | .45 | .056 |
| Site 2 | 2, 15 | 1.23 | .31 | .076 |
| Site 3 | 2, 13 | 3.16 | .06 | .196 |
| Site 4 | 2, 14 | .21 | .82 | .014 |
| Site 5 | 2, 25 | 2.14 | .13 | .079 |
| Site 6 | 2, 12 | 2.67 | .18 | .182 |

Data were analyzed using repeated-measures ANOVAs with Stimulus (CS+; CS+, CS-) as within-subjects factor.

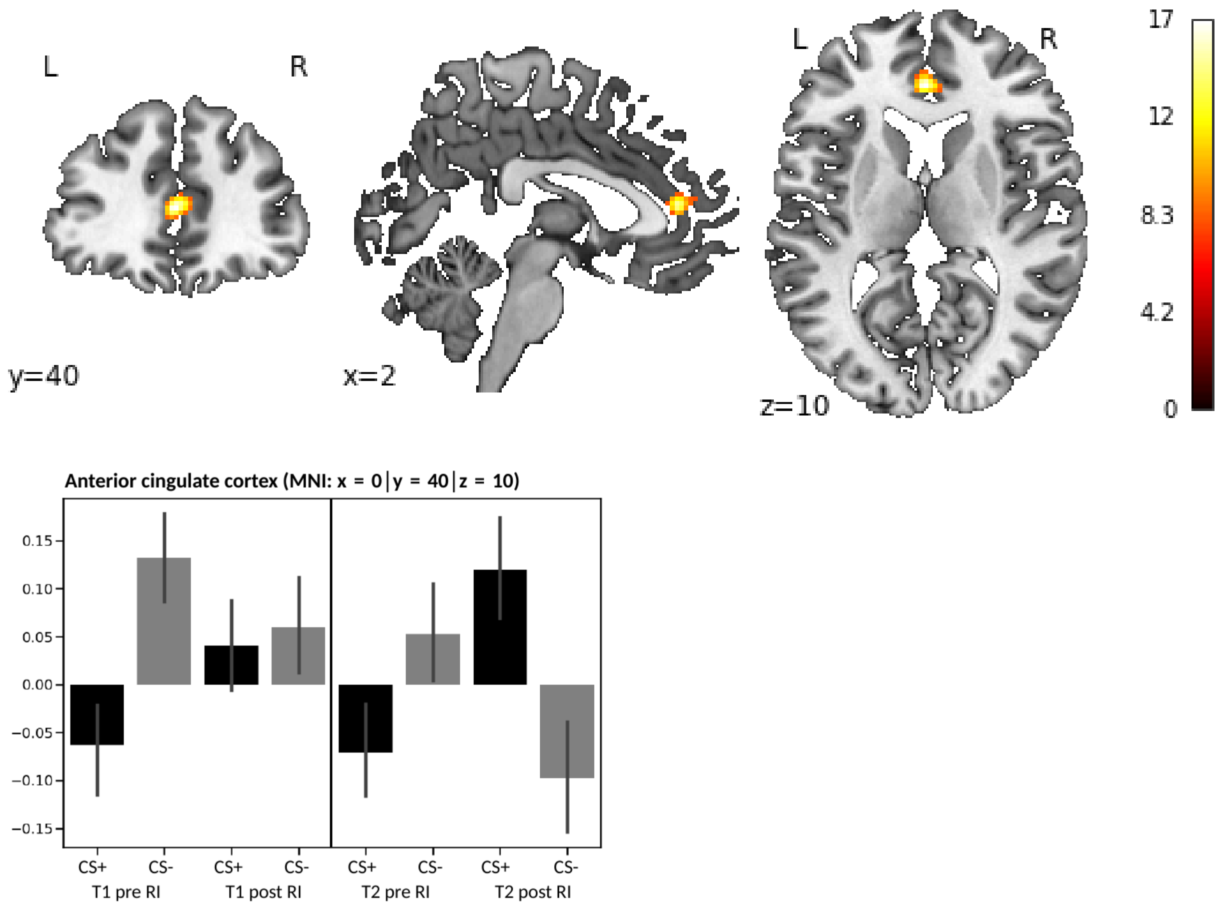


**Fig. S3.** Activation differences before and after reinstatement in the interaction CS-type X pre/post reinstatement. The contrast was calculated across both measurement points and assessed at *p*<.005 uncorrected with no cluster threshold.

**Table S4.** Conjuction (t1: Ex1 CS+ > CS-) ∩ (t2: Ex1 CS+ > CS-). All contrasts were assessed at *p*<.005 uncorrected with a cluster threshold of *k*=175.

| Region | hemi-sphere | no. voxels | MNI coordinates peak | | | | P-value FWE-corrected | |
|---|---|---|---|---|---|---|---|---|
| | | | x | y | z | t | peak | cluster |
| **(t1: Ex1 CS+ > CS-) ∩ (t2: Ex1 CS+ > CS-)** | | | | | | | | |
| Anterior insula + frontal + central operculum + caudate + pallidum | R | 3830 | 34 | 24 | 4 | 10.84 | <.001 | <.001 |
| Anterior insula + frontal operculum | L | 2341 | -30 | 20 | 8 | 9.41 | <.001 | <.001 |
| Superior frontal gyrus medial segment + middle cingulate gyrus + supplementary motor cortex | R | 4982 | 4 | 38 | 38 | 7.37 | <.001 | <.001 |
| Supramarginal gyrus + angular gyrus + parietal operculum | R | 2438 | 60 | -44 | 30 | 7.34 | <.001 | <.001 |
| Supramarginal gyrus + postcentral gyrus + parietal operculum | L | 1151 | -62 | -24 | 26 | 7.01 | <.001 | <.001 |
| Precentral gyrus + middle frontal gyrus | R | 736 | 46 | 2 | 46 | 5.71 | <.001 | .004 |
| Cerebellum | L | 413 | -16 | -76 | -28 | 4.68 | .042 | .066 |
| Middle + superior temporal gyrus | L | 229 | 52 | -28 | -6 | 4.67 | .043 | .374 |
| Middle + superior frontal gyrus | R | 542 | 38 | 50 | 16 | 4.42 | .114 | .021 |
| Middle cingulate gyrus + precentral gyrus medial segment | L | 310 | -2 | -22 | 30 | 4.14 | .297 | .175 |
| Middle frontal gyrus | L | 272 | -34 | 52 | 24 | 3.99 | .452 | .250 |
| **(t1: Ex1 CS- > CS+) ∩ (t2: Ex1 CS- > C+-)** | | | | | | | | |
| Precuneus + posterior cingulate gyrus + fusiform gyrus + hippocampus | | 2980 | 0 | -54 | 26 | 7.29 | <.001 | <.001 |
| Angular gyrus + superior parietal lobule | L | 3234 | -48 | -68 | 30 | 6.96 | <.001 | <.001 |
| Middle + superior temporal gyrus | L | 457 | -60 | -12 | -16 | 6.40 | <.001 | .045 |
| Medial frontal cortex + gyrus rectus + middle + superior frontal gyrus | R | 1898 | 2 | 40 | -18 | 6.37 | <.001 | <.001 |
| Angular gyrus + middle occipital gyrus + superior parietal lobule | R | 3198 | 44 | -62 | 32 | 5.68 | <.001 | <.001 |
| Hippocampus + fusiform gyrus + lingual gyrus | R | 756 | 32 | -40 | -8 | 5.36 | .002 | .004 |
| Pre- + postcentral gyrus medial segment | R | 2866 | 10 | -26 | 74 | 5.06 | .008 | <.001 |
| Pre- + postcentral gyrus | R | 1014 | 62 | -6 | 22 | 4.79 | .026 | .001 |

**Abbreviations:** CS+: conditioned stimulus that is followed by the unconditioned stimulus (US) with a reinforcement rate of 60% (only unpaired CS+ were included); CS-: conditioned stimulus that is never followed by an US; L: left; R: right; no. voxel: number of voxels per cluster; x, y, z: MNI coordinates.

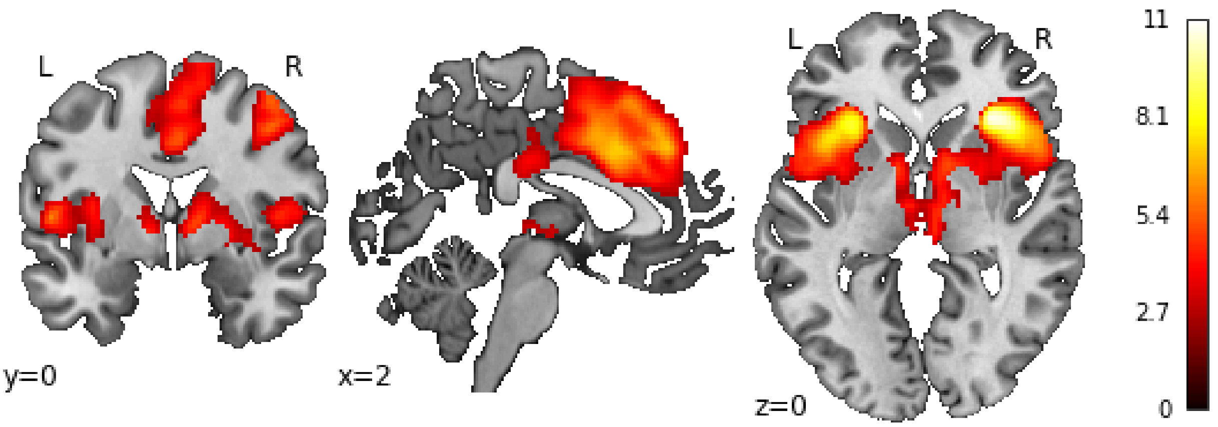


**Figure S4.** Conjuction (t1: Ex1 CS+ > CS-) ∩ (t2: Ex1 CS+ > CS-). All contrasts were assessed at $p$<.005 uncorrected with a cluster threshold of $k$=175.

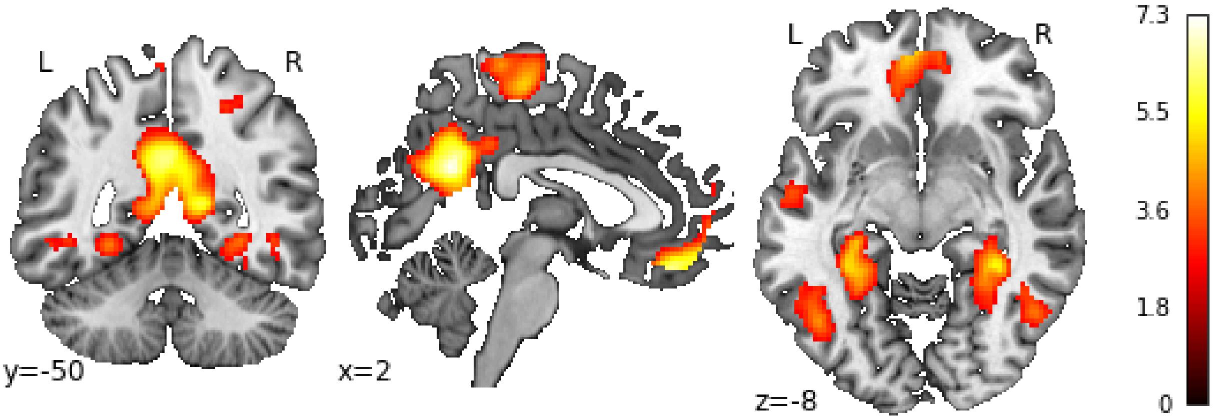


**Figure S5.** Conjuction (t1: Ex1 CS- > CS+) ∩ (t2: Ex1 CS- > CS+). All contrasts were assessed at $p$<.005 uncorrected with a cluster threshold of $k$=175.

**Table S5.** Activation difference CS+ vs. CS- at site 1 during first extinction phase across both measurement points. All contrasts were assessed at *p*<.005 uncorrected with a cluster threshold of *k*=175.

| Region | hemi-sphere | no. voxels | MNI coordinates peak | | | | P-value FWE-corrected | |
|---|---|---|---|---|---|---|---|---|
| | | | x | y | z | t | peak | cluster |
| **CS+ > CS-** | | | | | | | | |
| Anterior insula + frontal + central operculum | R | 1003 | 32 | 24 | 4 | 6.94 | <.001 | <.001 |
| Anterior insula + frontal operculum + putamen | L | 1468 | -30 | 18 | 8 | 6.43 | <.001 | <.001 |
| Supramarginal gyrus + angular gyrus+ parietal operculum | R | 803 | 58 | -42 | 32 | 5.12 | .018 | .001 |
| Superior frontal gyrus medial segment + middle cingulate gyrus | R | 890 | 6 | 34 | 38 | 4.80 | .064 | .001 |
| Supramarginal gyrus + parietal operculum + postcentral gyrus | L | 336 | -56 | -26 | 26 | 4.60 | .127 | .100 |
| Thalamus + pallidum + caudate | R | 249 | 10 | 0 | 4 | 3.92 | .734 | .257 |
| **CS- > CS+** | | | | | | | | |
| Medial frontal cortex + gyrus rectus + superior frontal gyrus | R | 4515 | 4 | 46 | -18 | 7.15 | <.001 | <.001 |
| Middle + superior temporal gyrus hippocampus | L | 2639 | -60 | -12 | -14 | 6.77 | <.001 | <.001 |
| Angular gyrus + middle temporal gyrus + fusiform gyrus + hippocampus | R | 4000 | 46 | -64 | 32 | 6.51 | <.001 | <.001 |
| Angular gyrus + middle occipital gyrus | L | 2391 | -46 | -68 | 30 | 6.23 | <.001 | <.001 |
| Precuneus + posterior cingulate gyrus | L | 1675 | -2 | -60 | 30 | 5.78 | .001 | <.001 |
| Precentral + postcentral gyrus | L | 6420 | -26 | -20 | 72 | 5.75 | .001 | <.001 |
| Thalamus | R | 257 | 16 | -32 | 6 | 3.93 | .725 | .235 |

**Abbreviations:** CS+: conditioned stimulus that is followed by the unconditioned stimulus (US) with a reinforcement rate of 60% (only unpaired CS+ were included); CS-: conditioned stimulus that is never followed by an US; L: left; R: right; no. voxel: number of voxels per cluster; x, y, z: MNI coordinates.

**Table S6.** Activation difference CS+ vs. CS- at site 2 during first extinction phase across both measurement points. All contrasts were assessed at *p*<.005 uncorrected with a cluster threshold of *k*=175.

| Region | hemi-sphere | no. voxels | MNI coordinates peak | | | | P-value FWE-corrected | |
|---|---|---|---|---|---|---|---|---|
| | | | x | y | z | t | peak | cluster |
| **CS+ > CS-** | | | | | | | | |
| Anterior insula + frontal + central operculum | R | 1268 | 36 | 26 | 2 | 5.83 | .001 | <.001 |
| Anterior insula + frontal + central operculum | L | 1101 | -40 | 14 | 2 | 5.19 | .011 | <.001 |
| Parietal operculum + supramarginal gyrus + postcentral gyrus | L | 395 | -56 | -28 | 24 | 5.11 | .016 | .080 |
| Supramarginal gyrus + parietal operculum + angular gyrus | R | 792 | 50 | -34 | 26 | 4.51 | .142 | .003 |
| Superior frontal gyrus medial segment + supplementary motor cortex + middle cingulate gyrus | R | 1035 | 2 | 36 | 40 | 4.39 | .214 | <.001 |
| Precentral gyrus + middle fronatl gyrus | R | 179 | 46 | 2 | 54 | 4.37 | .224 | .596 |
| **CS- > CS+** | | | | | | | | |
| Precuneus + posterior cingulate gyrus | R | 2122 | 8 | -52 | 14 | 6.02 | <.001 | <.001 |
| Middle + superior temporal gyrus | R | 349 | 62 | -6 | -18 | 4.77 | .058 | .124 |
| Angular gyrus + middle occipital gyrus | L | 1020 | -44 | -70 | 30 | 4.56 | .122 | <.001 |
| Middle + superior temporal gyrus | L | 410 | -58 | -12 | -16 | 4.52 | .136 | .069 |
| Angular gyrus + middle occipital gyrus | R | 959 | 50 | -64 | 32 | 4.49 | .155 | .001 |
| Postcentral + supramarginal gyrus | R | 410 | 36 | -24 | 42 | 4.03 | .544 | .069 |
| Inferior temporal gyrus | L | 194 | -46 | -50 | -8 | 4.02 | .560 | .528 |
| Hippocampus + parahippocampal gyrus + fusiform gyrus | R | 376 | 32 | -40 | -8 | 3.99 | .589 | .096 |

| | | | | | | | | |
|---|---|---|---|---|---|---|---|---|
| Hippocampus + parahippocampal gyrus + fusiform gyrus | L | 358 | -32 | -24 | -14 | 3.94 | .654 | .113 |
| Anterior cingulate gyrus + medial frontal gyrus + superior frontal gyrus medial segment | L | 463 | -10 | 34 | -8 | 3.75 | .849 | .042 |

**Abbreviations:** CS+: conditioned stimulus that is followed by the unconditioned stimulus (US) with a reinforcement rate of 60% (only unpaired CS+ were included); CS-: conditioned stimulus that is never followed by an US; L: left; R: right; no. voxel: number of voxels per cluster; x, y, z: MNI coordinates.

**Table S7.** Activation difference CS+ vs. CS- at site 3 during first extinction phase across both measurement points. All contrasts were assessed at *p*<.005 uncorrected with a cluster threshold of *k*=175.

| Region | hemi-sphere | no. voxels | MNI coordinates peak | | | | P-value FWE-corrected | |
|---|---|---|---|---|---|---|---|---|
| | | | x | y | z | t | peak | cluster |
| **<u>CS+ > CS-</u>** | | | | | | | | |
| Anterior insula + frontal operculum | R | 1339 | 30 | 26 | -6 | 7.71 | <.001 | <.001 |
| Superior frontal gyrus medial segment + anterior insula + thalamus | R | 7007 | 2 | 34 | 40 | 6.66 | <.001 | <.001 |
| Angular gyrus + supramarginal gyrus | R | 1002 | 60 | -46 | 30 | 5.41 | .005 | .001 |
| Cerebellum | L | 212 | -2 | -52 | -32 | 4.37 | .238 | .481 |
| Middle frontal gyrus | R | 498 | 50 | 16 | 46 | 4.35 | .256 | .036 |
| Supramarginal gyrus | L | 287 | -62 | -44 | 36 | 4.23 | .356 | .246 |
| **<u>CS- > CS+</u>** | | | | | | | | |
| Post- + precentral gyrus | L | 6560 | -8 | -36 | 70 | 5.43 | .005 | <.001 |
| Superior parietal lobule + inferior + middle temporal gyrus + superior occipital gyrus | R | 3688 | 28 | -66 | 36 | 5.16 | .014 | <.001 |
| Angular gyrus + occipital gyrus + superior parietal lobule | L | 1625 | -34 | -66 | 42 | 4.46 | .183 | <.001 |
| Hippocampus + parahippocampal gyrus + fusiform gyrus | L | 298 | -28 | -32 | -12 | 4.17 | .413 | .222 |
| Superior + middle temporal gyrus | R | 304 | 60 | -2 | -8 | 3.64 | .876 | .210 |
| Medial frontal cortex + gyrus rectus | R | 177 | 4 | 44 | -16 | 3.62 | .893 | .635 |

**Abbreviations:** CS+: conditioned stimulus that is followed by the unconditioned stimulus (US) with a reinforcement rate of 60% (only unpaired CS+ were included; CS-: conditioned stimulus that is never followed by an US; L: left; R: right; no. voxel: number of voxels per cluster; x, y, z: MNI coordinates.

**Table S8.** Activation difference CS+ vs. CS- at site 4 during first extinction phase across both measurement points. All contrasts were assessed at *p*<.005 uncorrected with a cluster threshold of *k*=175.

| Region | hemi-sphere | no. voxels | MNI coordinates peak | | | | P-value FWE-corrected | |
|---|---|---|---|---|---|---|---|---|
| | | | x | y | z | t | peak | cluster |
| **<u>CS+ > CS-</u>** | | | | | | | | |
| Frontal + central operculum + anterior insula + putamen | R | 2966 | 38 | 26 | 2 | 7.21 | <.001 | <.001 |
| Supplementary motor + middle cingulate gyrus | R | 4245 | 4 | 22 | 54 | 5.65 | .002 | <.001 |
| Frontal operculum + anterior insula | L | 1976 | -44 | 20 | 8 | 5.26 | .010 | <.001 |
| Parietal operculum + supramarginal gyrus | R | 798 | 46 | -26 | 26 | 4.95 | .036 | .001 |
| Supramarginal gyrus + parietal operculum | L | 261 | -58 | -26 | 24 | 4.85 | .052 | .213 |
| Precentral gyrus + middle frontal gyrus | R | 311 | 46 | 4 | 48 | 4.48 | .187 | .122 |
| Middle frontal gyrus | R | 360 | 30 | 44 | 18 | 4.42 | .226 | .071 |
| Middle + superior temporal gyrus | R | 235 | 58 | -22 | -8 | 4.22 | .397 | .284 |
| Brain stem + thalamus | R | 216 | 8 | 28 | -8 | 4.04 | .602 | .349 |
| Middle cingulate gyrus | R | 223 | 4 | -20 | 40 | 3.89 | .774 | .323 |
| **<u>CS- > CS+</u>** | | | | | | | | |

| | | | | | | | | |
|---|---|---|---|---|---|---|---|---|
| Superior parietal lobule + occipital gyrus | R | 3074 | 26 | -68 | 42 | 5.21 | .013 | <.001 |
| Precentral gyrus medial segment | | 2200 | 0 | -26 | 68 | 4.62 | .118 | <.001 |
| Angular gyrus + middle occipital gyrus | L | 1711 | -40 | -68 | 28 | 4.55 | .149 | <.001 |
| Inferior temporal gyrus | R | 504 | 48 | -60 | -6 | 4.27 | .352 | .016 |
| Precuneus + posterior cingulate gyrus | L | 907 | -4 | -54 | 32 | 4.19 | .437 | <.001 |
| Hippocampus + parahippocampal gyrus + fusiform gyrus | R | 376 | 34 | -30 | -14 | 4.18 | .448 | .060 |
| Superior + middle frontal gyrus | L | 565 | -18 | 28 | 44 | 4.10 | .531 | .009 |
| Medial frontal cortex + gyrus rectus + anterior cingulate gyrus | L | 462 | -6 | 50 | -10 | 4.09 | .544 | .024 |
| Postcentral gyrus | R | 200 | 62 | -8 | 38 | 4.05 | .590 | .413 |

**Abbreviations:** CS+: conditioned stimulus that is followed by the unconditioned stimulus (US) with a reinforcement rate of 60% (only unpaired CS+ were included); CS-: conditioned stimulus that is never followed by an US; L: left; R: right; no. voxel: number of voxels per cluster; x, y, z: MNI coordinates.

**Table S9.** Activation difference CS+ vs. CS- at site 5 during first extinction phase across both measurement points. All contrasts were assessed at *p*<.005 uncorrected with a cluster threshold of *k*=175.

| **Region** | **hemis-phere** | **no. voxels** | **MNI coordinates peak** | | | | **P-value FWE-corrected** | |
|---|---|---|---|---|---|---|---|---|
| | | | **x** | **y** | **z** | **t** | **peak** | **cluster** |
| **CS+ > CS-** | | | | | | | | |
| Frontal operculum + anterior insula + opercular part of the inferior frontal gyrus | R | 2579 | 38 | 24 | 6 | 7.55 | <.001 | <.001 |
| Anterior insula + frontal operculum + opercular part of the inferior frontal gyrus | L | 1544 | -30 | 20 | 8 | 6.20 | <.001 | <.001 |
| Superior frontal gyrus medial segment + supplementary motor cortex + middle cingulate gyrus | R | 2969 | 6 | 34 | 40 | 5.74 | .001 | <.001 |
| Supramarginal gyrus + angular gyrus + parietal operculum + middle + superior temporal gyrus | R | 2206 | 58 | -42 | 28 | 5.72 | .001 | <.001 |
| Supramarginal gyrus + postcentral gyrus | L | 1039 | -62 | -26 | 24 | 5.42 | .003 | <.001 |
| Precentral gyrus + middle frontal gyrus | R | 423 | 42 | 4 | 46 | 5.40 | .004 | .034 |
| Cerebellum + fusiform gyrus | L | 344 | -28 | -52 | -26 | 5.07 | .015 | .080 |
| Thalamus + brain stem | R | 436 | 6 | -22 | 0 | 4.52 | .131 | .030 |
| Middle + superior temporal gyrus | R | 215 | 56 | -28 | -6 | 3.98 | .619 | .341 |
| Middle + superior frontal gyrus | R | 304 | 32 | 54 | 24 | 3.92 | .690 | .125 |
| **CS- > CS+** | | | | | | | | |
| Angular gyrus + posterior cingulate gyrus + precuneus | L | 4457 | -38 | -68 | 34 | 5.53 | .002 | <.001 |
| Middle + inferior frontal gyrus + precentral gyrus | L | 447 | -46 | 24 | 24 | 5.15 | .011 | .026 |
| Middle + inferior temporal gyrus | L | 251 | -52 | -60 | -8 | 4.83 | .040 | .228 |
| Superior + middle occipital gyrus | R | 1472 | 28 | -82 | 36 | 4.67 | .075 | <.001 |
| Superior frontal gyrus medial segment + medial frontal cortex | L | 1363 | -14 | 46 | 0 | 4.57 | .110 | <.001 |
| Superior + middle frontal gyrus | L | 369 | -22 | 22 | 50 | 4.52 | .134 | .061 |
| Precentral gyrus medial segment + postcentral gyrus medial segment | R | 631 | 8 | -24 | 62 | 4.36 | .222 | .004 |
| Central operculum + post- + precentral gyurs | L | 510 | -60 | -8 | 12 | 4.35 | .232 | .014 |
| Pre- + postcentral gyrus | R | 444 | 52 | -2 | 26 | 4.01 | .579 | .027 |
| Postcentral + precentral gyrus | R | 333 | 48 | -20 | 58 | 3.89 | .716 | .091 |
| Hippocmapus + parahippocampal gyrus + fusiform gyrus + lingual gyrus | L | 386 | -30 | -30 | -16 | 3.80 | .820 | .051 |
| Fusiform gyrus + lingual gyrus | R | 204 | 34 | -42 | -6 | 3.77 | .846 | .384 |
| Precentral + postcentral gyrus | L | 522 | -38 | -16 | 64 | 3.71 | .891 | .012 |

**Abbreviations:** CS+: conditioned stimulus that is followed by the unconditioned stimulus (US) with a reinforcement rate of 60% (only unpaired CS+ were included); CS-: conditioned stimulus that is never followed by an US; L: left; R: right; no. voxel: number of voxels per cluster; x, y, z: MNI coordinates.

**Table S10.** Activation difference CS+ vs. CS- at site 6 during first extinction phase across both measurement points. All contrasts were assessed at *p*<.005 uncorrected with a cluster threshold of *k*=175.

| Region | hemi-sphere | no. voxels | MNI coordinates peak | | | | P-value FWE-corrected | |
|---|---|---|---|---|---|---|---|---|
| | | | x | y | z | t | peak | cluster |
| **CS+ > CS-** | | | | | | | | |
| Frontal operculum + anterior insula | R | 2138 | 40 | 20 | 2 | 5.45 | .004 | <.001 |
| Postcentral gyrus + supramarginal gyrus | L | 1656 | -58 | -22 | 28 | 5.35 | .006 | <.001 |
| Middle cingulate gyrus + supplementary motor cortex | R | 3720 | 4 | 6 | 40 | 5.15 | .014 | <.001 |
| Central + parietal operculum + supramarginal gyrus | R | 2177 | 48 | -18 | 22 | 5.02 | .023 | <.001 |
| Central + frontal operculum + anterior insula | L | 1763 | -46 | 6 | 0 | 4.97 | .028 | <.001 |
| Superior parietal lobule + postcentral gyrus + precuneus | R | 314 | 20 | -46 | 66 | 4.00 | .576 | .213 |
| Cerebellum | L | 524 | -20 | -56 | -48 | 3.99 | .587 | .035 |
| Precentral gyrus + middle frontal gyrus | R | 303 | 44 | -2 | 58 | 3.98 | .596 | .235 |
| Precuneus | L | 227 | -12 | -72 | 36 | 3.85 | .743 | .451 |
| Middle frontal gyrus | L | 412 | -32 | 54 | 28 | 3.65 | .913 | .090 |
| Precentral gyrus medial segment + middle cingulate gyrus | L | 199 | -14 | -30 | 44 | 3.58 | .946 | .561 |
| Superior frontal gyrus + middle frontal gyrus | R | 321 | 24 | 62 | 20 | 3.36 | .995 | .040 |
| **CS- > CS+** | | | | | | | | |
| Posterior cingulate cortex + precuneus | L | 535 | -8 | -52 | 22 | 4.81 | .051 | .032 |
| Angular gyrus | L | 192 | -36 | -62 | 28 | 3.86 | .728 | .591 |
| Medial frontal cortex + gyrus rectus | R | 212 | 2 | 40 | -16 | 3.75 | .836 | .508 |

**Abbreviations:** CS+: conditioned stimulus that is followed by the unconditioned stimulus (US) with a reinforcement rate of 60% (only unpaired CS+ were included); CS-: conditioned stimulus that is never followed by an US; L: left; R: right; no. voxel: number of voxels per cluster; x, y, z: MNI coordinates.

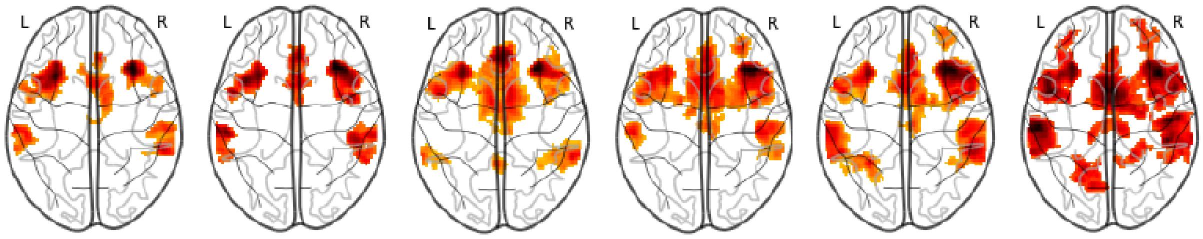


**Figure S6.** Activation difference CS+ > CS- during first extinction phase across both measurement points. From left to right site 1 to 6. For each site a separate 2nd -level-analysis was performed. All contrasts were assessed at *p*<.005 uncorrected with a cluster threshold of *k*=175.

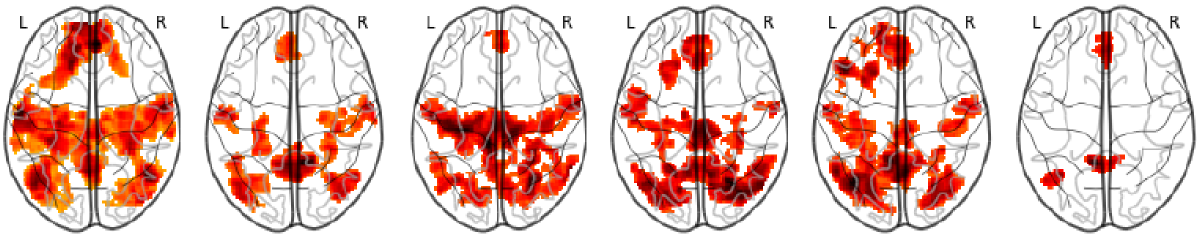


**Figure S7.** Activation difference CS- > CS+ during first extinction phase across both measurement points. From left to right site 1 to 6. For each site a separate 2nd-level-analysis was performed. All contrasts were assessed at *p*<.005 uncorrected with a cluster threshold of *k*=175.
